# Continuous Infusion of the CXCR4 Antagonist Plerixafor for WHIM Syndrome

**DOI:** 10.1101/2025.04.19.25325865

**Authors:** David H. McDermott, Shamik Majumdar, Daniel Velez, Elena Cho, Zhanzhou Li, Ji-Liang Gao, Megan C. Grieco, Monica G. Lawrence, Susana L. Silva, Leslie A. Castelo-Soccio, Dean Follmann, Philip M. Murphy

## Abstract

WHIM (Warts, Hypogammaglobulinemia, Infections and Myelokathexis) syndrome is an ultrarare inborn error of immunity caused by heterozygous, gain-of-function *CXCR4* mutations that impede leukocyte egress from bone marrow, resulting in panleukopenia. The CXCR4 antagonist plerixafor (AMD3100, Mozobil) durably reverses panleukopenia and in most WHIM patients induces wart regression; however, its short half-life requires twice daily injection. To develop a simpler, cheaper and potentially more effective method of drug delivery, we conducted a Phase 1 study of WHIM patients given plerixafor 0.02-0.08 mg/kg/d by continuous subcutaneous infusion using an OmniPod insulin pump, and assessed compliance as well as effects on blood leukocyte counts, infections, chronic skin conditions and adverse events. Six patients were treated for a total of 6.3 patient-years; one patient dropped out early for personal reasons. The drug infusion rate was adjusted to achieve a normal absolute lymphocyte count and an absolute neutrophil count >500 cells/µl in all patients. An average of 2.1 infections/patient-year occurred (range 0-4). Treatment of two infections involved brief hospitalization. On plerixafor, partial wart regression occurred in 3 of 4 patients, a single molluscum contagiosum infection regressed and a chronic post-Mohs surgical wound epithelialized. There were 3 serious adverse events, but none was attributable to the treatment. All patients preferred pump administration over syringe injection. Thus, in WHIM patients a continuous infusion pump may be a convenient, safe and potentially cost-effective means of delivering plerixafor chronically to correct panleukopenia and to improve chronic skin conditions. Clinicaltrials.gov NCT00967785.

## Introduction

WHIM syndrome is an ultrarare combined primary immunodeficiency disorder. Manifestations include warts, hypogammaglobulinemia and acute barrier site infections that vary in severity and frequency and are caused by common bacteria and viruses.^1-7^ Although rarely life-threatening, recurrent infection may lead to chronic sequelae, particularly bronchiectasis and hearing loss. Moreover, patients may have increased HPV- and EBV-associated cancer risk.^7,8^

The genetic cause of WHIM syndrome is heterozygous, gain-of-function mutation of the panleukocyte chemokine receptor CXCR4.^9-11^ This causes myelokathexis (myeloid hyperplasia, dysmorphic bone marrow neutrophils and severe neutropenia), resulting from an exaggeration of two normal CXCR4-dependent functions: nascent neutrophil retention in bone marrow and senescent neutrophil homing from blood to bone marrow.^5^ Hypogammaglobulinemia is incompletely penetrant.^12^

Other leukocyte functions appear normal, and infection may reverse neutropenia,^13^ perhaps explaining low mortality. Consequently, bone marrow allotransplantation risk may exceed benefit.^14^ Immunoglobulin replacement and G-CSF are common treatments;^5^ however, trials of safety and efficacy are lacking, and both treatments are expensive, inconvenient and fail to reverse lymphopenia and monocytopenia. Moreover, chronic G-CSF risks myelodysplasia and acute myeloid leukemia, and commonly induces bone pain, arthralgia and non-compliance; with high doses myelofibrosis and bone marrow failure may occur.^15,16^

To address these limitations, two small molecule CXCR4 antagonists have been advanced: the parenteral agent plerixafor (Mozobil, AMD3100; Sanofi-Genzyme) and the oral agent mavorixafor (Xolremdi; X4 Pharmaceuticals), which both recently completed Phase 3 trials.^17,18^ Both drugs were well-tolerated and durably increased neutrophil and lymphocyte counts. Compared with placebo, mavorixafor reduced annualized infection rate, but not infection severity or warts, and is now FDA-approved for WHIM syndrome.^18^ Compared with G-CSF, plerixafor was not superior for infection severity; partial and sometimes complete wart regression occurred only on plerixafor.^17^ Plerixafor (combined with G-CSF) was FDA-approved in 2009 for hematopoietic stem cell (HSC) mobilization. Its patent expired in 2023, spawning generic versions and >85% decline in market price. WHIM patients may access it off-label, and through Sanofi’s Patient Access Program.^16^

Like G-CSF, plerixafor comes in single use vials. For clinical research in chronic disease applications, plerixafor cost is amplified by the requirements for syringe compounding under GMP conditions and twice daily injections are given owing to the short half-life.^16,17,19^ To further reduce cost, simplify treatment and improve receptor coverage and potentially clinical outcome, we performed a pilot, single center, investigator-initiated, open label, Phase 1 trial of continuous, subcutaneous plerixafor infusion in WHIM syndrome.

## Methods

### Study subjects

Subjects ages 18-75 with severe neutropenia and a carboxy-terminal *CXCR4* pathogenic variant were eligible. Patients on supplemental immunoglobulin and/or prophylactic antibiotics could continue these treatments. All subjects signed written informed consent documents approved by the NIH Institutional Review Board (Clinicaltrials.gov NCT00967785). The study complied with the Declaration of Helsinki. An independent safety monitoring committee monitored safety. All authors had access to primary data which were analyzed by DHM, SM, DF, MG and PMM.

### Drug delivery system

Sanofi-Genzyme (Cambridge, MA) provided plerixafor by agreement with NIAID/NIH. Vials containing 20 mg/ml plerixafor in 1.2 ml sterile physiologic saline were stored at room temperature in light-protected boxes in the Investigational Drug Management Section of the NIH Clinical Center (NIH CC) Pharmacy. Drug was administered by OmniPod pump (Insulet Corporation, Acton, MA) at the same total daily dose range as our previous trials using twice-daily syringes, 0.02-0.08 mg/kg/d.^16,17,19^ This pump had previously been successfully used off-label for a trial of Parathyroid Hormone 1-34.^20^ Plerixafor stability and biocompatibility with the OmniPod system was assessed by HPLC and mass spectroscopy (Supplemental Appendix Methods).

### Preclinical testing of continuous plerixafor delivery

Preclinical experiments used male littermate C57BL/6 mice kept in specific pathogen-free conditions under NIAID Animal Care and Use Committee-approved protocol LMI-8E. Wild-type (WT) mice were from Jackson Laboratories (Bar Harbor, ME). WHIM model mice have been previously described and were provided by Drs. Balabanian and Bachelerie (INSERM, Paris).^21^ Plerixafor was infused continuously at 3 µg/hr in ∼11-13-week old WT and WHIM model mice using subcutaneously implanted Alzet 2006 osmotic minipumps (Durect Corporation, Cuppertino, CA).^21^ Pumps were primed with phosphate buffered saline (PBS) at 37°C for 60 h then filled with 200 µl of either PBS or 20 mg/ml AMD3100 (Sigma-Aldrich, St. Louis, MO) in PBS. The infusion rate was 0.15 µl/hr for both AMD3100 and PBS. Blood was sampled for leukocyte phenotyping one day before infusion and at one-week intervals for 6 weeks, as described.^22,23^

### Clinical trial of continuous plerixafor delivery

Patients were admitted to the NIH CC and trained to control the Omnipod using a personal diabetes manager (PDM), which adjusts rate in increments of 5 µl/hour and delivers drug for 72 hours before signaling the need for replacement. Leukocyte-mobilizing drugs were stopped at least 2 days before the day baseline blood leukocyte counts were determined, defined as day -1 (relative to starting plerixafor) (Figure 1). On day -1 a saline-filled Pod was applied, and a saline bolus given followed by continuous saline infusion for 24 hours at the equivalent rate necessary to deliver 0.03 mg/kg/day of plerixafor. Blood was collected for leukocyte quantitation just before and ∼3, 6, 9, 12 and 24 hours into the saline infusion. Immediately after the 24-hour sampling timepoint (start of day 0) a single 0.015 mg/kg actual body weight plerixafor bolus was administered from a plerixafor-loaded Pod, and serial blood leukocyte counts were determined at ∼3, 6, 9, 12 and 24 hours afterwards. This dose is 6.25% of the FDA-approved 0.24 mg/kg dose for HSC mobilization. On day +1, continuous plerixafor infusion began at 0.03 mg/kg/day monitoring leukocyte counts at ∼3, 6, 9, 12 and 24 hours thereafter and then daily until discharge on day +3 or +4. This daily dose is 12.5% of the FDA-approved 0.24 mg/kg bolus dose for HSC mobilization. Leukocyte counts were monitored at home or NIH then at least ∼every 3 months with Pod changes every 3 days for 1-2 years. Dose was adjusted from 0.02-0.08 mg/kg/day to optimize ANC and ALC and was recorded by the PDM.

**Figure 1.**
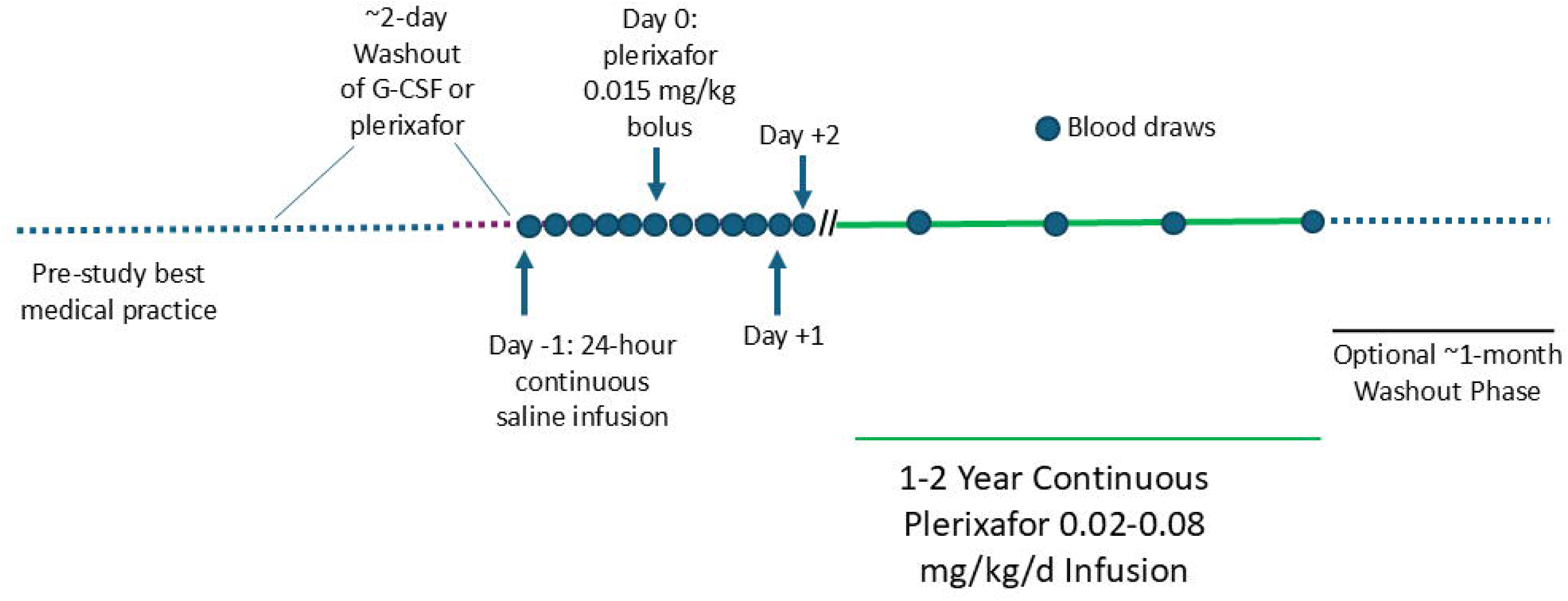
Design of a Phase 1/2 clinical trial of chronic continuously infused plerixafor treatment of WHIM patients. The schema indicates study phases, treatments and blood draws. Study phase durations are not drawn to scale.

An optional ∼30-day washout phase could follow the study with additional blood counts before resuming pre-study treatment. The infusion rate was adjusted to maintain an ANC>=500 cells/µl. If ANC exceeded 4000 cells/µl without signs of infection, the rate was reduced. Drug failure requiring discontinuation was defined in two ways: 1) if by month 3 the ANC repeatedly remained <250 neutrophils/µl at the maximal allowed dose; or 2) if the average value of all available ANCs on drug remained <250 neutrophils/µl or <2-fold greater than the baseline average pre-drug value.

### Patient assessments

In addition to complete blood counts with differential, blood chemistries were monitored at baseline and at a minimum at months 2 & 3, then ∼every 3 months at NIH or local laboratories, as described.^17^ Immunoglobulin levels, lymphocyte subset levels, neutrophil chemokine receptor expression and patient photography were obtained at baseline and the ∼6-month and ∼12-month NIH visits. Descriptions of infections and adverse events were obtained from patients and local providers.^17^

### Flow cytometry

Neutrophil surface chemokine receptor expression was determined by flow cytometry using the patient and healthy donor blood sampling timepoints, gating strategy, monoclonal antibodies and staining methods detailed in the Supplemental Methods and Supplemental Figures S1 and S2.

### Statistical analysis plan

Analysis of the safety primary endpoint involved a yes/no determination of drug-related Grade III or IV toxicities. Safety and side effects were assessed from the safety tests mentioned previously and case report forms. Analysis of the efficacy primary endpoint was also a yes/no determination based on an average ANC >250/µl over the entire treatment period.

As a secondary endpoint, we compared the range (maximum less the minimum) of all leukocyte counts obtained during the 24-hour period following the plerixafor bolus and during the first 24-hour period of continuous plerixafor. A paired t-test was used to compare the difference in ranges on bolus vs continuous plerixafor.

To assess long term performance of continuous infusion of plerixafor, we described leukocyte counts over time. We also descriptively assessed how warts changed over time by comparing photographs at baseline, at ∼month 6 on plerixafor and at the end of the infusion, defining a 50% decline in individual wart areas between baseline and the end of study as significant. We also compared leukocyte subtype levels at baseline and at the end of treatment. A paired t-test was used for this comparison. If a patient failed to complete the maximum period of treatment because of voluntary withdrawal, their data were still used in the analysis. Therefore, the analysis was not on an “Intention to Treat” basis but rather an actual treatment basis. Tests were two-sided with a type I error rate of 0.05.

## Results

### Preclinical validation of continuous plerixafor infusion for WHIM syndrome

We first tested plerixafor compatibility with the OmniPod system. Compared to the external reference, static incubation of plerixafor in a Pod at 37°C for 72 hours generated no new peaks or shifts in retention time by HPLC, and no new mass spectrum peaks in the mass/charge (m/z) range between 120 and 530 Daltons (data not shown). Drug concentrations in the pump after incubation and in the source vial were both ∼20 mg/ml, validating stability in the Pod.

Next, preclinical experiments were performed using an Alzet osmotic minipump. All mice tolerated pump implantation and total white blood cell counts durably increased over the 6-week observation period after infusing AMD3100 at 3 μg/hr (Figure 2A and B). WT and WHIM model mice responded similarly (Figure 2A).

**Figure 2.**
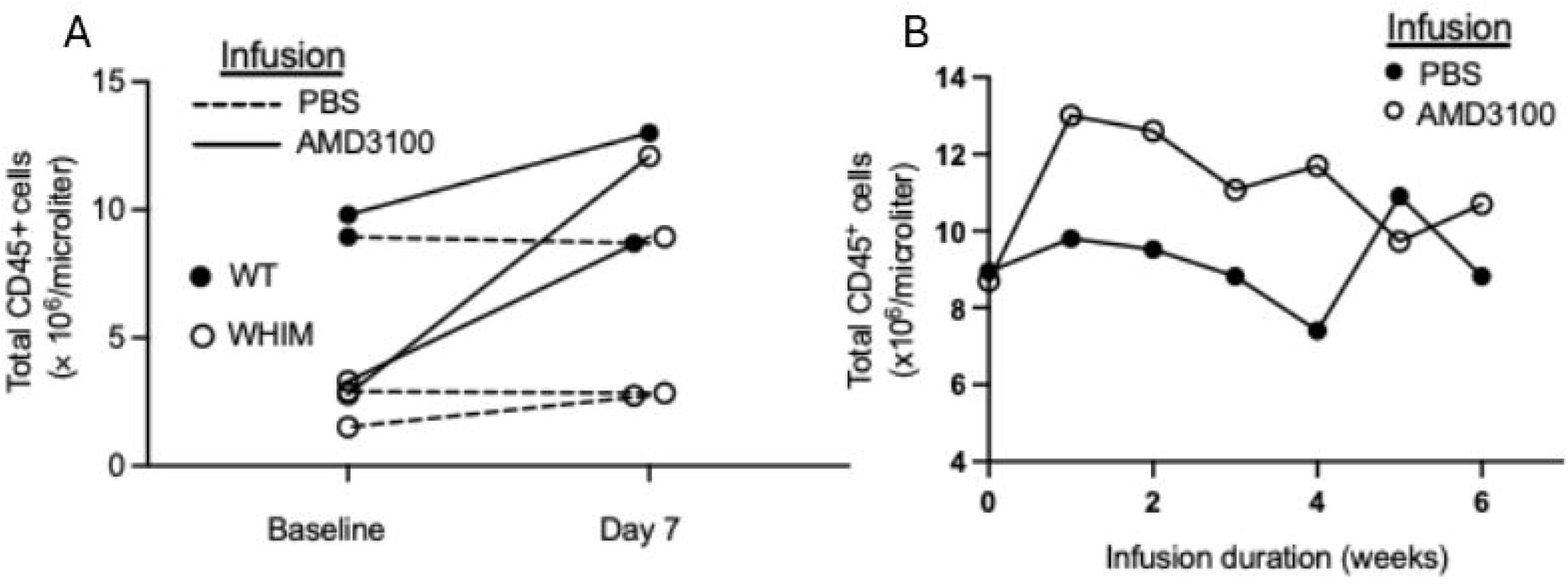
Continuous infusion of AMD3100 in mice durably increases circulating leukocyte counts. A) Wild type and WHIM model C57BL/6 mice treated for one week. B) Wild type mice treated for 6 weeks. Each symbol denotes an individual mouse. Infusions and genotypes are coded in the insets. Data are from a single experiment.

### Characteristics of continuous infusion clinical trial participants

We then enrolled six WHIM patients for continuous plerixafor infusion (Table 1). All patients had a history of recurrent infections (Supplemental Table 1). Patients P1-P5 had bronchiectasis;^17^ P1 had received supplemental oxygen for >15 years. P2 and P3, who had been splenectomized 13 and 23 years before enrollment, respectively, and P1 and P6 were all receiving supplemental immunoglobulin.

**Table 1:** Characteristics of WHIM patients treated with continuously infused plerixafor.

| Patient | Sex | Race | Age range (yrs)<sup>&</sup> | CXCR4 mutation | WHIM Phenotypes | Time since splenectomy (yrs) | Prior Treatment History<sup>@</sup> |  |  |  |  |
| --- | --- | --- | --- | --- | --- | --- | --- | --- | --- | --- | --- |
|  |  |  |  |  |  |  | IgT (yrs) | Proph. Abx | Plerixafor by BiD SQ injection (mos) | Mavorixafor (mos) | G-CSF (~total yrs) |
| P1 | F | W | 48-57 | R334* | whim | na | 36 | Yes | 20 | 0 | 30 |
| P2 | M | W | 38-47 | R334* | whim | 13 | 14 | No | 0 | 0 | 1 |
| P3 | M | W | 18-27 | S324Vfs20* | him | 23 | 8 | Yes | 78 | 0 | 21 |
| P4 | M | H | 18-27 | R334* | whim<sup>#</sup> | na | 0 | Yes | 14 | 0 | 12 |
| P5 | F | W | 58-67 | R334* | wim | na | 0 | No | 14 | 16 | episodic |
| P6 | M | W/NA | 18-27 | R334* | whim | na | 14 | No | 2 | 0 | 14 |

Abbreviations: W=White, H=Hispanic, NA=Native American; mos, months; yrs, years; WHIM phenotype: w=warts, h=hypogammaglobulinemia, i=infections, m=myelokathexis; IgT=time on supplemental immunoglobulin treatment; Proph. Abx, prophylactic antibiotics (continued during study); BiD SQ, twice daily subcutaneous; na, not applicable.
<sup>&</sup>Ages are given within 10-year intervals for privacy. Age eligibility was 18-75.
<sup>#</sup>warts (w) in this patient completely regressed 5 years before entry during prior treatment with plerixafor.<sup>17</sup>
<sup>@</sup>Duration of prior plerixafor treatment is tabulated as the number of months. Treated was ended before enrollment in the present study by at least 3 years in all subjects except P3 who stopped twice daily injections 1 week before enrollment.

All 6 patients were G-CSF-experienced, and all but P3 and P6 were receiving it up until starting the study. P3 was G-CSF-intolerant, as reported previously,^16^ and had been treated for 7 years with twice daily plerixafor injections before enrollment. P1, P4, P5 and P6 were also plerixafor-experienced from participation in our Phase 3 study of twice daily subcutaneous injection, all ending >3 years before the present study.^16,17,19^ Plerixafor treatment successfully mobilized leukocytes in P1, P4 and P5 in the Phase 3 study, defined as a durable ANC rise to >500 cells/µl and of ALC to >1000 cells/µl. P6 had failed plerixafor at month 3 during Phase 3 when ANC stayed below the prespecified threshold of 500 cells/µl.

P5 had received open label mavorixafor for ∼8 months following participation as a placebo control subject in the Phase 3 mavorixafor trial,^18^ but had stopped treatment 1 month before enrollment in the present study because of a rising creatinine. Creatinine was stable in the interim and had been in the normal range during Phase 3 twice daily plerixafor injections.^17^

P1 began continuous infusion in September 2020 and the final patient, P6, ended infusion in June 2023 (Supplemental Figure 1). Four patients received continuous plerixafor for one year. P3 was treated for 2 years. P6 dropped out after 1.5 months, after taking a new job. Compliance, as documented by the PDM, was 94 +/- 3% of intended dose, range 83-99%.

### Leukocyte mobilization by continuous plerixafor infusion in WHIM patients

After G-CSF or plerixafor washout, all day -1 baseline ANC, ALC and AMC values were low except for the ALC of P2, which was normal (Figure 3A). Asplenia is unlikely to explain this anomaly since P3 and other asplenic WHIM patients have low ALC levels typical of WHIM patients with an intact spleen.

**Figure 3.**
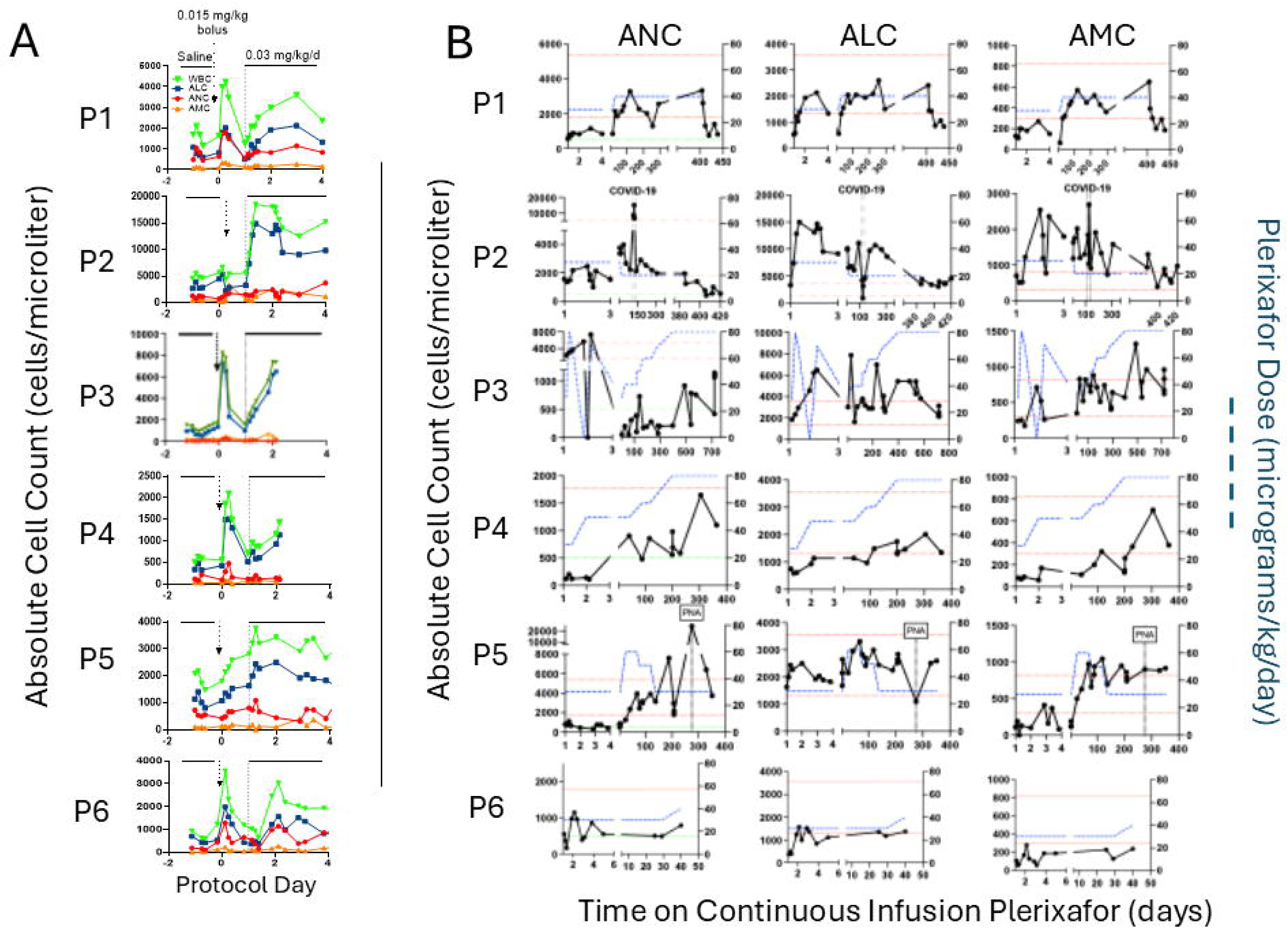
Continuous infusion of plerixafor durably increases complete blood counts in WHIM patients. A) Time course for week 1 of the protocol for each patient. Serial blood counts are shown after the washout (baseline day -1 values), and during 24-hour intervals of saline infusion, after plerixafor bolus (given at day 0, vertical arrow) and during continuous plerixafor infusion starting at day +1 (dashed vertical line), as indicated at the top of the figure. The cell types are color-coded in the inset of the top panel. Each graph corresponds to the patient indicated to its left. B) Time course for continuous plerixafor infusion for each patient. Each row of graphs corresponds to the patient indicated to its left. Each column of graphs corresponds to the leukocyte subtype at the top. Horizontal dashed red lines demarcate the normal range for each leukocyte subtype. The horizontal dashed lines on the ANC graphs demarcate 500 cells/µl. Plerixafor doses and leukocyte counts are quantitated on the right and left y-axes, respectively. The time and duration of COVID-19 and pneumococcal pneumonia (PNA) infections are demarcated by pairs of vertical dashed lines for patients P2 and P5, respectively.

During 24-hour saline infusion, we observed minor transient increases in absolute counts in all patients. Bolus plerixafor injection on day zero by Omnipod induced an expected rapid and large increase in total white blood cell count in P1 and P3-P6 (Figure 3A). However, the subtype bolus responsiveness rank order was highly variable: ALC∼ANC>AMC for P1 and P6, ALC>>>>ANC∼AMC for P3, and ALC>>ANC>>AMC for P4 and P5. Again, P2 was an outlier in that both ANC and AMC increased but inexplicably ALC did not, despite increasing during continuous plerixafor infusion after day +1. All increases were transient except for those of P5, which were unexpectedly sustained at peak throughout the 24-hour timepoint after bolus injection. The plerixafor bolus dose was sufficient to bring the ALC peak into or above the normal range for all 6 patients. However, peak ANC was normal at this dose only for P1 and P2, and peak AMC was normal only for P1, P2 and P3.

On continuous infusion, all patients met the primary efficacy definition of success: an average ANC >250/µL over the treatment period (Figure 3A and B). Thus, the estimated proportion of efficacy was 1.00, with a Clopper-Pearson confidence interval of 0.541-1.000. One-sample t-test results on the difference in log10 ranges for white blood cell counts after the bolus plerixafor injection versus during the first 24 hours of continuous plerixafor infusion showed no significant differences (p=0.148 for ANC and p=0.853 for ALC) (Figure 3A). P6’s ALC reached the normal range, and his ANC exceeded 500 cells/µl by 1.5 months when he dropped out. ALC and AMC normalized for all other patients. ANC normalized for P1, P2 and P5, and exceeded 500 cells/µl for P3 and P4. Only P4 required an infusion rate increase for ALC to normalize, and P2’s rate was decreased once owing to an undesirably high initial ALC response, aligning with his anomalously high baseline ALC. P2’s ANC normalized on the starting rate of 0.03 mg/kg/d, whereas the rate was increased once to normalize ANC for P1 and P5. For P3 and P4, the infusion rate was increased five and three times, respectively, to attain an ANC >500 cells/µl, but neither patient normalized ANC even at the maximum allowed dose. This is consistent with their previously reported relative ANC resistance to augmentation by twice daily plerixafor injection.^16,17^

### Chemokine receptor expression on WHIM neutrophils

We next analyzed surface expression of the chemokine receptors CXCR1, CXCR2 and CXCR4 on circulating neutrophils before and after starting plerixafor (Supplemental Figures 1 and 2, Figure 4A). At the day 0 baseline measurement after pre-treatment washout, the patients had much lower proportions of neutrophils expressing all three receptors compared to healthy donors (HD). With one outlier, baseline variance for CXCR4^+^ patient neutrophils was low, in contrast to highly variable neutrophil mobilization efficiency by plerixafor. During treatment, the proportion of neutrophils expressing CXCR1 and CXCR2 increased in most patients (Fig. 4B), whereas CXCR4 values markedly decreased at the 6-month timepoint, consistent with plerixafor blocking CXCR4-antibody binding (Fig. 4B).^24^ Results were variable at the one-year timepoint.

**Figure 4.**
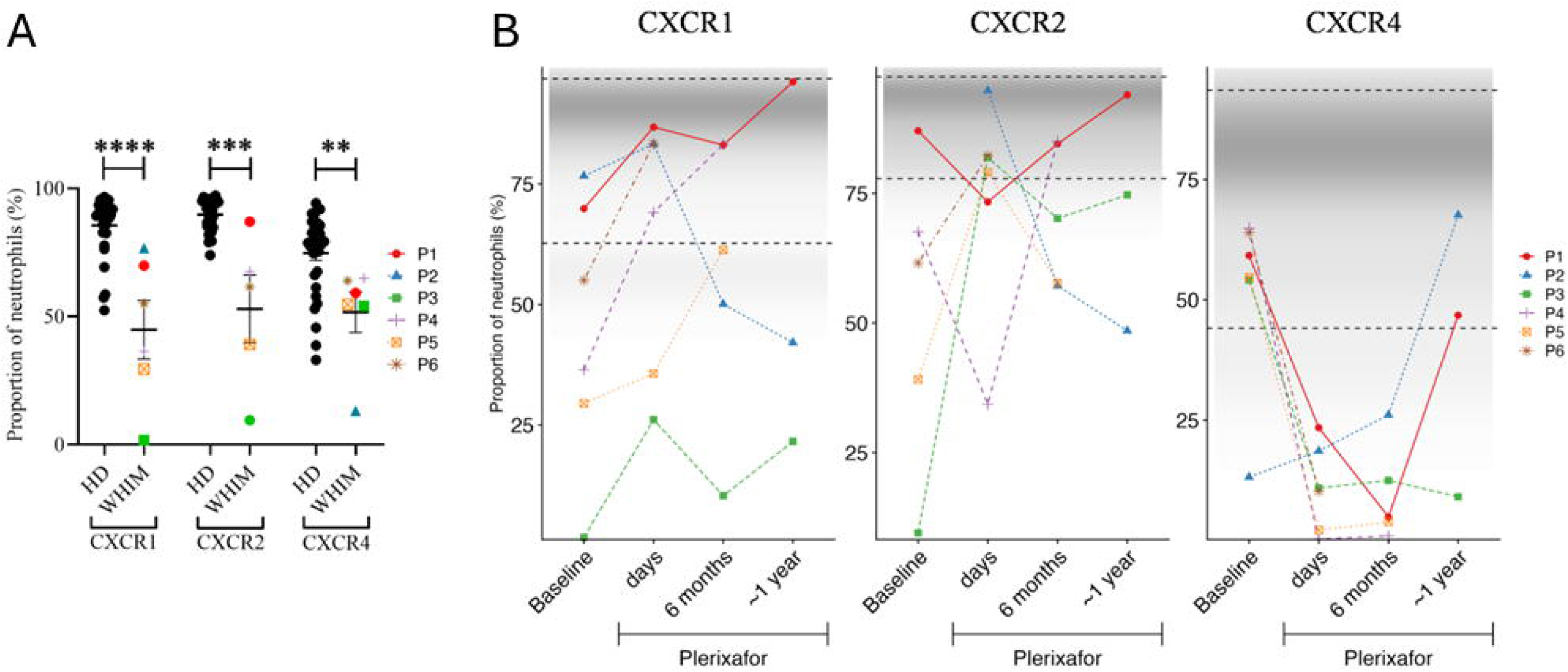
WHIM patients at baseline possess a high proportion of neutrophils deficient in CXCR1, CXCR2 and CXCR4 that is increased in some patients by chronic continuous infusion of plerixafor. A) Baseline. Comparison of neutrophil chemokine receptor expression in healthy donors and WHIM study subjects on d0 before plerixafor. Statistical significance was calculated using the Mann–Whitney U test; **, p<0.01; ***, p<0.001; ****, p<0.0001. HD, healthy donor; WHIM, WHIM patients. B) Time course on plerixafor. ‘days’ refers to measurement of neutrophils obtained 3-6 days after the baseline timepoint. The distribution of values for healthy donors is conveyed graphically by the density of the shaded region in each graph. Horizontal dashed lines demarcate 2 standard deviations from the mean value for 33 healthy donors aggregated from the indicated timepoints. The chemokine receptor corresponding to each graph is indicated at the top. Data in both panels are individual values corresponding to the patient color-coded at the right.

### Mobilization of lymphocyte subtypes by continuous plerixafor infusion in WHIM patients

ALC normalization by plerixafor reflected augmentation of baseline levels of total B cells and CD4 and CD8 positive T cells (Figure 5A). NK cell levels were normal for 4 patients at baseline and were not significantly augmented by plerixafor. Asplenic patients P2 and P3 were exceptional in having normal baseline total B cell levels. Memory B cell levels normalized for P1-P3 and P5, but not P4. Nevertheless, baseline immunoglobulin levels, which varied among patients, were unaffected by treatment (Figure 5B). Asplenic patient P3 had normal baseline IgA and IgM levels, whereas both levels were low for asplenic patient P2. The 4 patients with spleens were all severely B lymphopenic at baseline but had diverse immunoglobulin levels.

**Figure 5.**
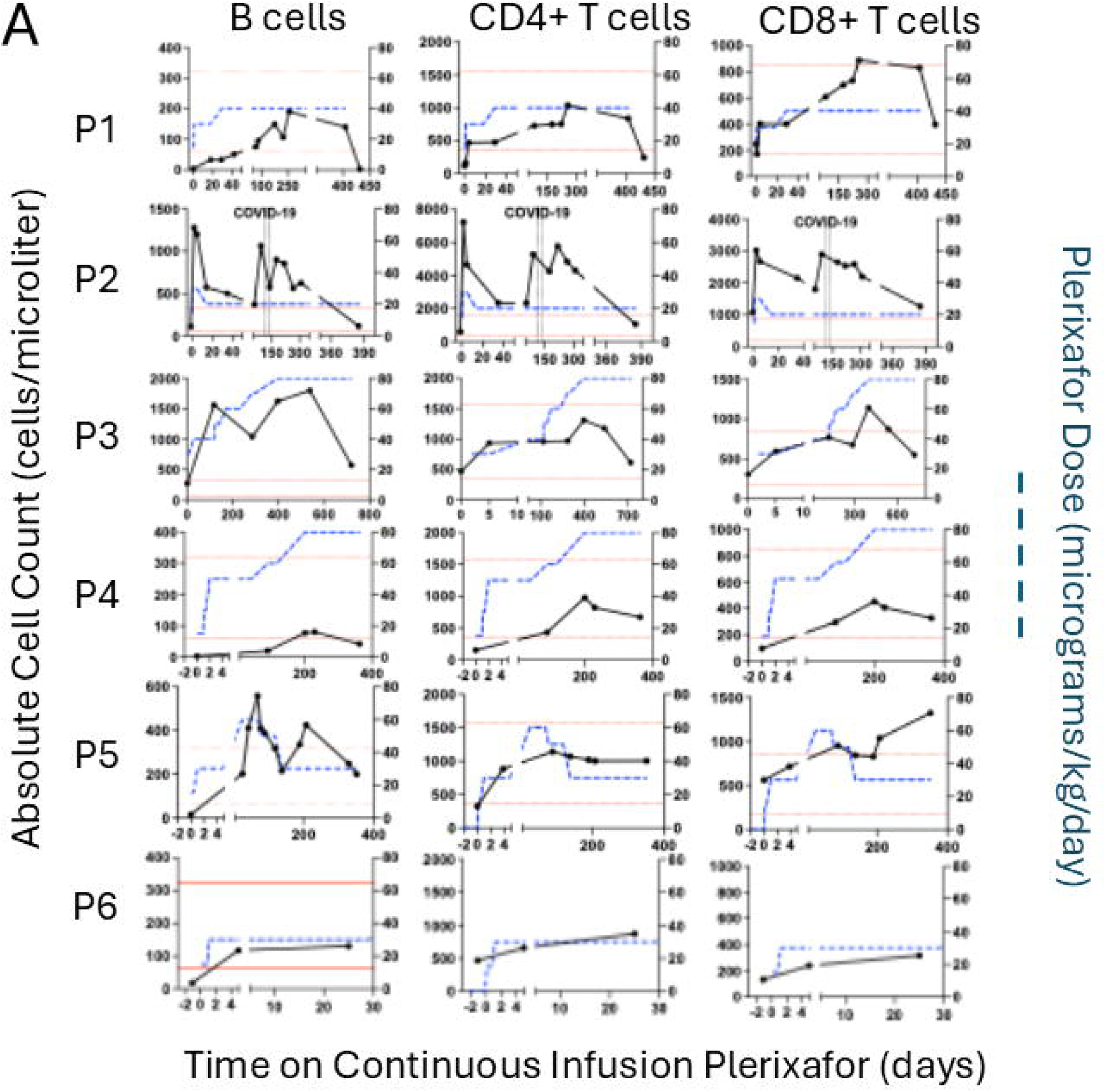
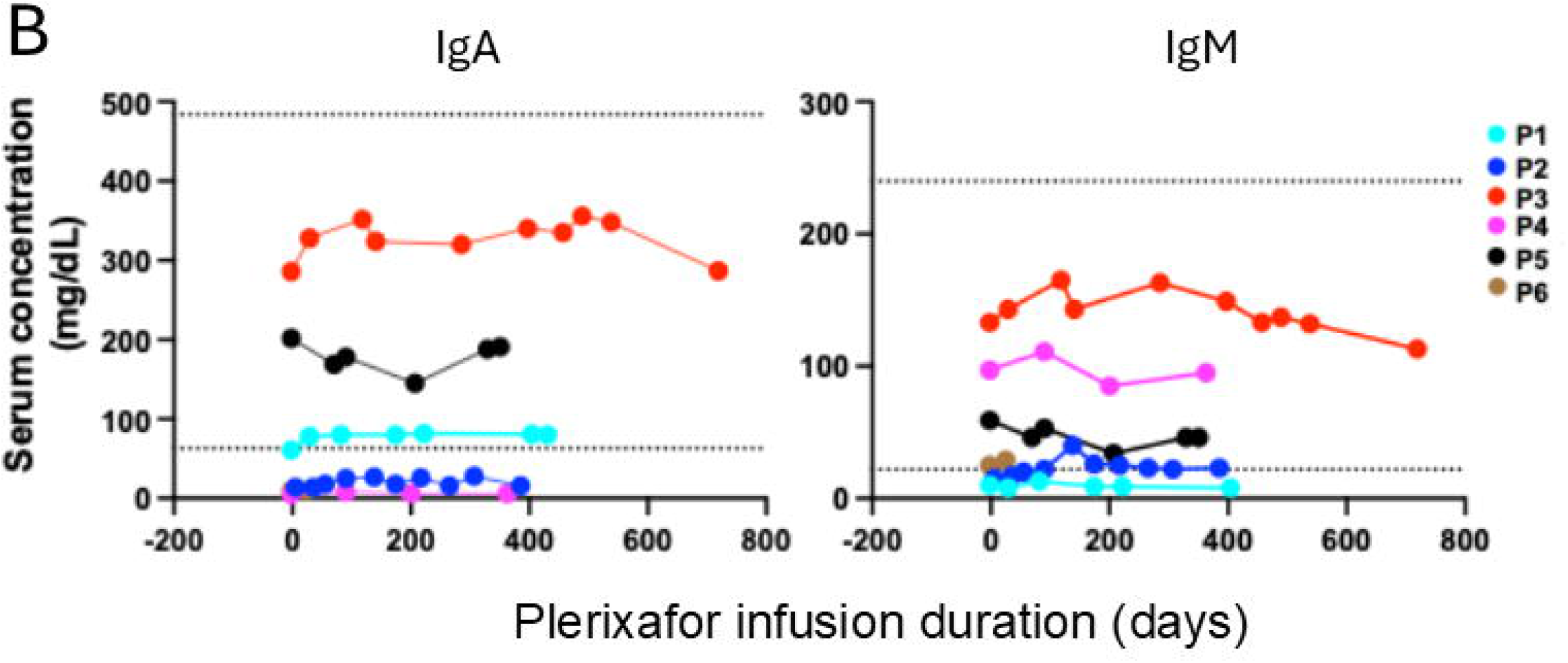
Continuous infusion of plerixafor durably increases circulating lymphocyte subtypes but not immunoglobulin levels in WHIM patients. A) Lymphocyte subtypes. Each row of graphs corresponds to the patient indicated to its left. Each column of graphs corresponds to the lymphocyte subtype indicated above the column. Plerixafor doses and lymphocyte counts are quantitated on the right and left y-axes, respectively. The time and duration of COVID-19 and pneumococcal pneumonia (PNA) infections are demarcated by pairs of vertical dashed lines for patients P2 and P5, respectively. Horizontal dotted red lines demarcate the normal range for adults at the NIH Clinical Center Department of Laboratory Medicine for each subtype. B) Serum immunoglobulin levels. Each graph corresponds to the immunoglobulin type indicated at the top. Horizontal dotted black lines demarcate the normal range for each parameter established for adults by the NIH-CC Department of Laboratory Medicine. IgG data are not shown because most patients were receiving supplemental immunoglobulin. In both panels, each symbol represents a single value for a single patient color-coded in the figure.

### Regression of chronic skin conditions in WHIM patients during continuous plerixafor infusion

Five patients had chronic skin infections: 4 had warts and one had *Molluscum contagiosum* infection. During treatment, no new warts appeared, and existing warts did not enlarge. Three of the 4 patients with warts (P1, P2 and P5) experienced partial wart regression after treatment (Table 2, Supplemental Table 2, Figure 6A). P2 and P5 also applied imiquimod (P2 from month 8-9 to hand and genital warts and P5 from months 0-2 and 6-10, applied 3 times per week to the right hand). P5 had a major wart burden on both hands and legs that had not regressed previously during twice daily plerixafor and imiquimod therapy during Phase 3 and had persisted thereafter. A large left leg flat wart area regressed during continuous plerixafor infusion. P1 had partial wart regression on previous Phase 1 and 3 plerixafor trials and continued to reduce several wart areas on her fingers during the present study. P2 had partial wart regression on both hands and wrists. P3 and P4 were wart-free at baseline, and Patient 6 had multiple wart areas that did not regress during his 1.5 months treatment. P4, who previously had a complete response of a large forehead flat wart area on twice-daily plerixafor injections during Phase 3, had no new warts but developed multiple umbilicated pubic lesions caused by *Molluscum contagiosum* virus prior to the present study (Figure 6B), which almost completely resolved by month-12 of continuous plerixafor. He applied imiquimod to some of the lesions from month 7-9.

**Figure 6.**
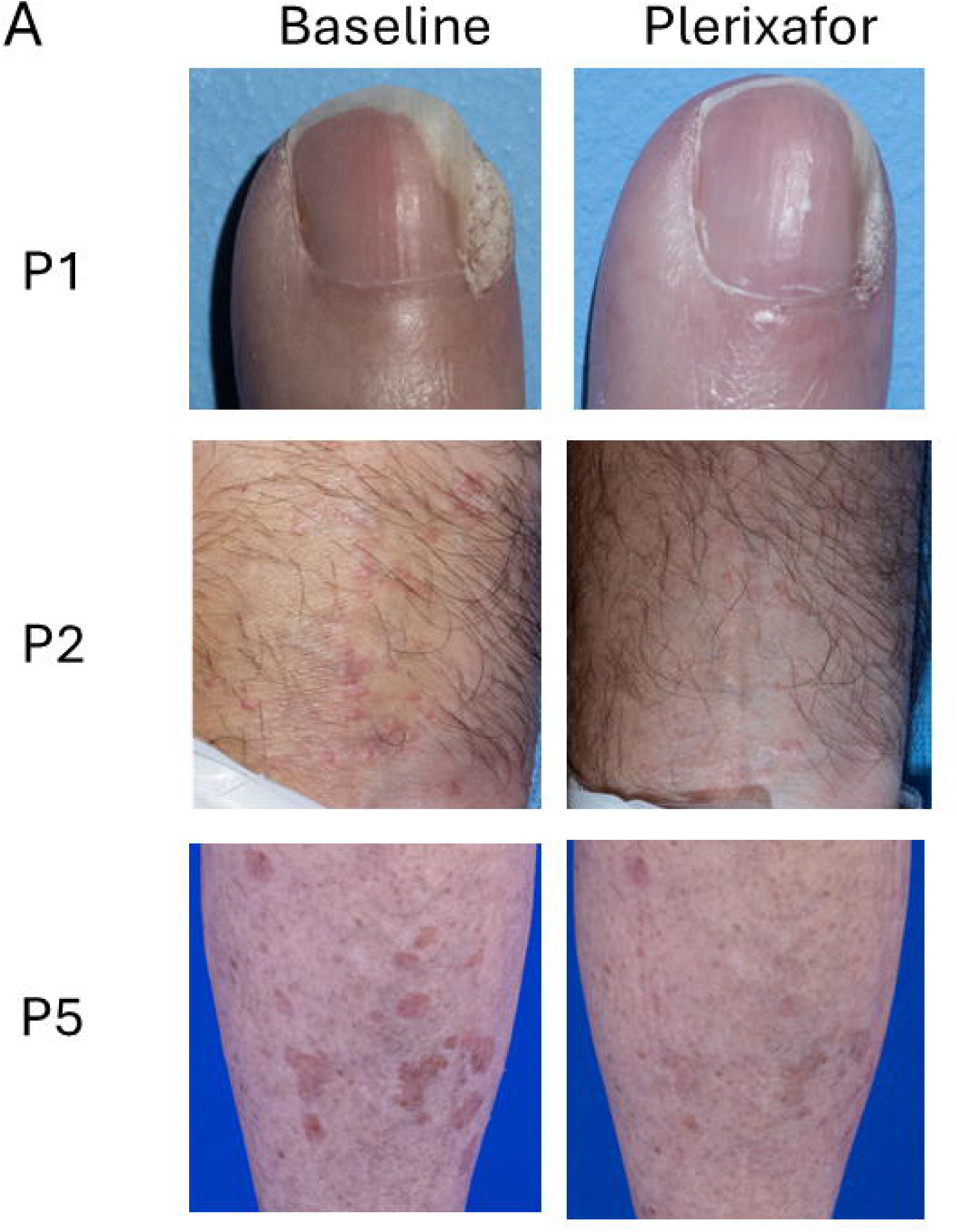
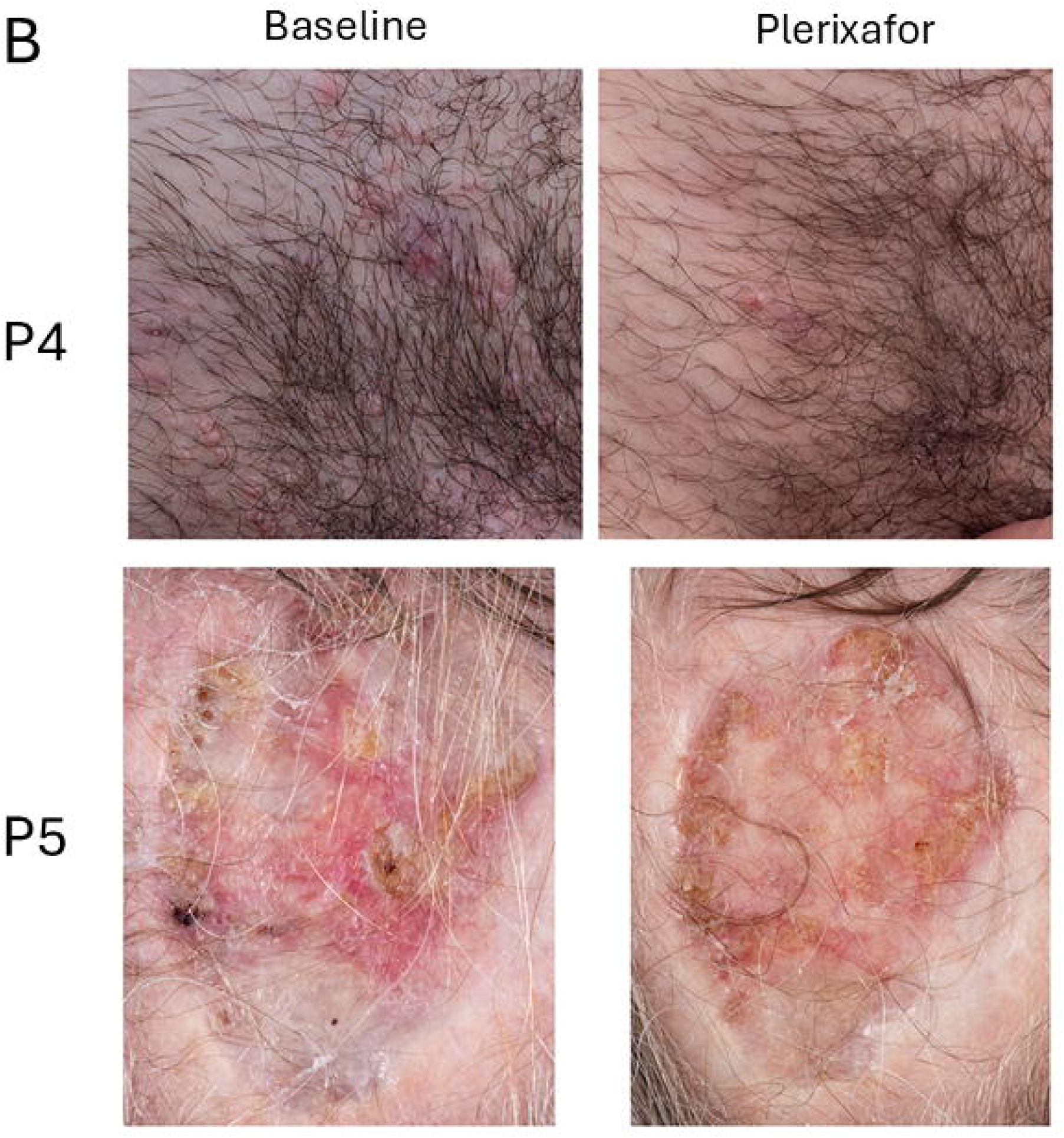
Improvement of skin conditions during continuous infusion of plerixafor in WHIM patients. Images at baseline (left) and from the end of plerixafor treatment (right) are shown for the patients identified to the left of the corresponding row. A) Wart areas. B) Molluscum contagiosum (P4) and chronic cranial defect post Mohs surgery for basal cell carcinoma (P5). Comprehensive assessments of wart changes on drug are detailed in Table 2 and Supplemental Table 2.

**Table 2.** Wart status at baseline and responses during continuous infusion of plerixafor in WHIM patients. A partial response was defined as >50% regression of a discrete wart area.

|  | **Patient Total, *n*** |
| --- | --- |
| **Patients on study** | 6 |
| **Wart status at randomization** |  |
| Positive history of warts | 5 |
| Warts present at baseline of study | 4 |
| **Wart responses during plerixafor infusion** |  |
| Partial | 3 |
| Complete | 0 |
| Increased burden | 0 |
| No improvement | 1<sup>a</sup> |

Abbreviations: *n*, number
<sup>a</sup>Patient P6 withdrew from the study after 1.5 months for social reasons.

P5 had an uninfected, chronic, 3X5 cm scalp wound with exposed bone from prior Mohs surgery for basal cell carcinoma that had persisted for >10 years. Skin grafting failed in 2016. During continuous plerixafor infusion and wound care, we observed extensive epithelialization of this lesion (Figure 6B).

### Acute infections in WHIM patients during plerixafor infusion

Thirteen acute, non-fatal infections occurred on plerixafor (range, 0-4/patient; average, 2.1/patient-year), all but one at skin, airway or gastrointestinal barrier sites. Two were serious adverse events requiring hospitalization and intravenous therapy, but not intensive care--COVID-19 for P2 and a *S. pneumoniae* pneumonia/bacteremia complicated by pulmonary embolus for P5 (Table 3). ANC rose from the plerixafor-supported baseline at the onset of these infections, then returned to the plerixafor-supported baseline after recovery, with reciprocal changes for ALC (Figure 3B).

**Table 3:**
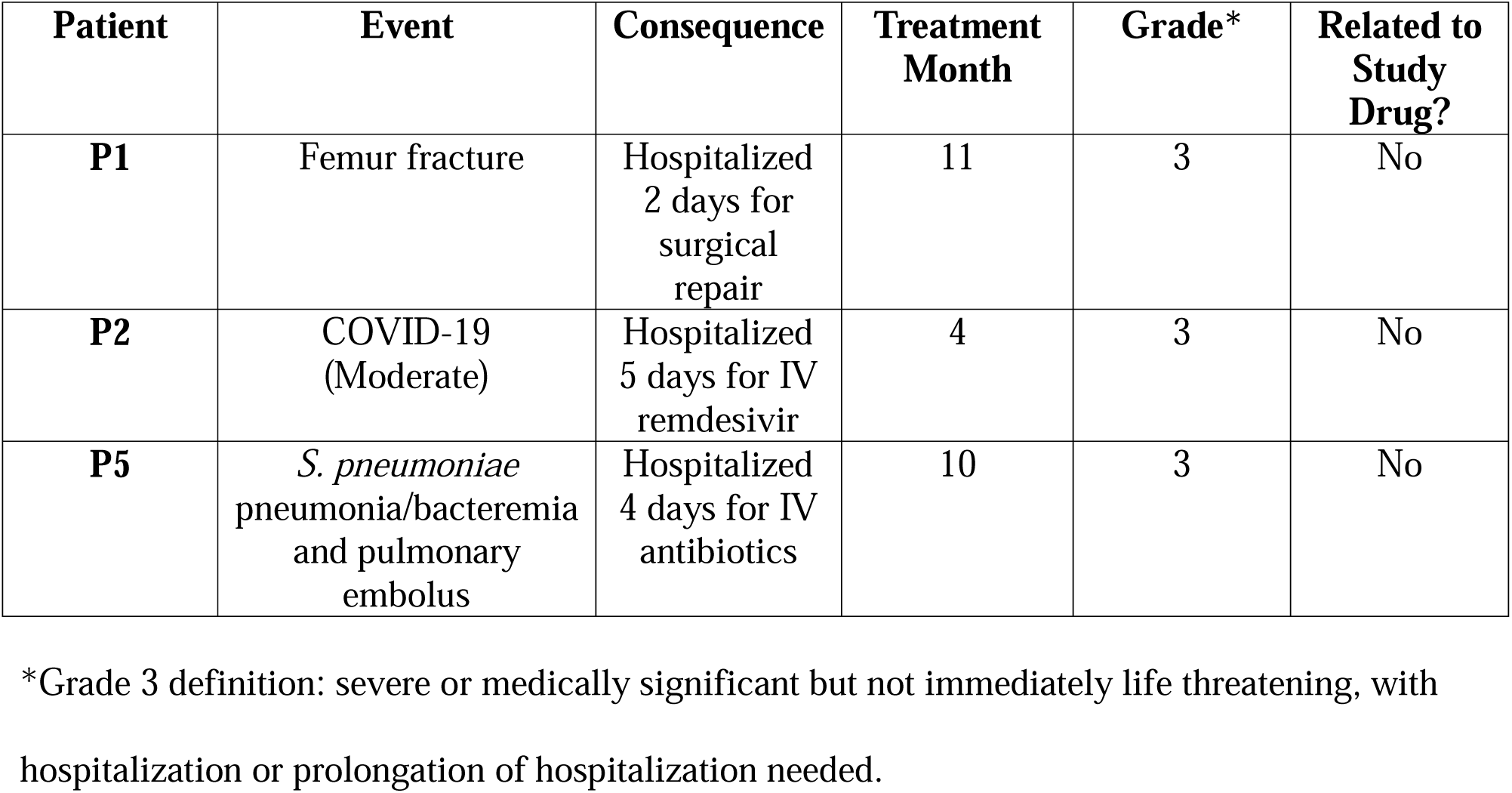
Serious Adverse Events during continuously infused plerixafor treatment of WHIM patients.

A causative organism was defined in 2 other infections, *Clostridium difficile* colitis in P5, diagnosed 3 weeks after completing treatment with Augmentin, Ciprofloxacin and Metronidazole for new onset diverticulitis, and Group A *Streptococcus* pharyngitis in P2 (Table 4). Two minor cases of cellulitis at the Pod cannula site were judged probably related to the study treatment. Both cases resolved during a short course of oral amoxicillin-clavulanate, and after scheduled Pod replacement at a new location. There were three cases of gastroenteritis. Two occurred in P1, both treated with hydration. The third, in P3, was treated with Cefuroxime and Metronidazole. There were three upper respiratory tract infections, all treated with an oral antibiotic, and one skin abscess, treated with Cephalexin. The courses of antibiotics were given for 9.2 +/- 1.2 days (range 5 to 16 days).

**Table 4:** Acute infections occurring during continuous plerixafor infusion of WHIM patients.

| Patient | Infections | Organism | Grade* | Onset (month on study) | Related to Study Drug? | Treatment Route (duration in days) | Outcome |
| --- | --- | --- | --- | --- | --- | --- | --- |
| **P1** | URTI | nd | 2 | 1 | No | Oral (5) | Resolved |
|  | Gastroenteritis | nd | 1 | 3 | No | None | Resolved |
|  | Gastroenteritis | nd | 1 | 8 | No | None | Resolved |
| **P2** | Pharyngitis | *Streptococcus* (rapid test) | 2 | 2 | No | IM/oral (1/14) | Resolved |
|  | COVID-19 (Moderate) | SARS-CoV-2 | 3 | 4 | No | IV (5) | Resolved |
| **P3** | Gastroenteritis | nd | 2 | 1 | No | Oral (8) | Resolved |
|  | Cannula-site cellulitis | nd | 2 | 5 | Yes | Oral (7) | Resolved |
|  | Cannula-site cellulitis | nd | 2 | 7 | Yes | Oral (7) | Resolved |
| **P4** | Skin Abscess | nd | 2 | 4 | No | Oral (10) | Resolved |
| **P5** | URTI | nd | 2 | 4 | No | Oral (7) | Resolved |
|  | Diverticulitis | nd | 2 | 5 | No | Oral (16) | Resolved |
|  | Colitis | *Clostridium difficile* | 2 | 6 | No | Oral (10) | Resolved |
|  | Pneumonia/bacteremia | *Streptococcus pneumoniae* | 3 | 10 | No | IV/oral (4/9) | Resolved |
| **P6** | None | na | na | na | na | na | na |

Abbreviations: URTI, Upper Respiratory Tract Infection; IM, intramuscular; IV, intravenous; nd, not determined; na, not applicable
\*Definitions--Grade 1: mild or asymptomatic, no intervention needed; Grade 2: moderate with minimal intervention needed; Grade 3: severe or medically significant but not immediately life threatening, with hospitalization or prolongation of hospitalization.

### Adverse events in WHIM patients during plerixafor infusion

Three Grade III toxicities occurred, unrelated to treatment. Two were the hospitalized infections described above. The third was a hip fracture from a fall by P1, who underwent orthopedic surgery, without adjusting the plerixafor dose, and healed at the expected rate. Thus, the proportion of subjects having a serious adverse event was 0.5, with a Clopper-Pearson confidence interval of 0.118-0.882.

Other non-infectious adverse events were minor (Table 5). Two were judged related to the treatment. The first, mild eczematous dermatitis, occurred in 3 patients, principally on the hands and feet, beginning ∼one month after starting plerixafor and lasting up to 6 months. This AE was also observed during twice daily plerixafor injections in Phase 3.^17^ The plerixafor infusion rate was not changed, and the problem resolved either without treatment or on moisturizers or topical corticosteroids.

**Table 5:** Non-infectious adverse events during continuous plerixafor infusion of WHIM patients.

| Patient | Adverse Event | Treatment<br>(duration in<br>days) | Time of onset<br>(month) | Grade* | Related to<br>treatment? |
| --- | --- | --- | --- | --- | --- |
| **P1** | Dermatitis | Emollient | 1 | 1 | Yes |
|  | Femur fracture | Surgery | 11 | 3 | No |
| **P2** | Dermatitis | Topical<br>corticosteroid | 1 | 2 | Yes |
| **P3** | None | na | na | na | na |
| **P4** | Conjunctivitis | None | 7 | 1 | No |
| **P5** | GERD | stop verapamil | 1 | 1 | No |
|  | Alopecia | stop verapamil | 1 | 1 | No |
|  | Eosinophilia/ABPA | Posaconazole<br>(90) | 5 | 2 | Yes |
|  | Anemia | None | 10 | 1 | No |
|  | Pulmonary embolus | Apixaban (180) | 10 | 3 | No |
| **P6** | Dermatitis | None | 1 | 1 | Yes |

Abbreviations: GERD, gastroesophageal reflux disease; ABPA, allergic bronchopulmonary aspergillosis; na, not applicable
\*Grade definitions: 1, mild or asymptomatic, no intervention needed; 2, moderate with minimal intervention needed; 3, severe or medically significant but not immediately life threatening, with hospitalization or prolongation of hospitalization.

The second occurred in P5, who had baseline bronchiectasis and chronic allergic bronchopulmonary aspergillosis, involving chronic colonization with *Aspergillus fumigatus*, mild eosinophilia and an elevated IgE level, including a positive *Aspergillus* precipitans test. Six months after starting plerixafor, her eosinophil count increased, peaking at ∼4500 cells/µl, without a change in chronic cough or chest CT. Plerixafor infusion rate was reduced, and posaconazole was given, with subsequent conversion of her sputum culture to negative for *Aspergillus fumigatus* and decline in her eosinophil counts.

### Patient preferences

All patients preferred continuous Omnipod delivery for plerixafor over daily injections of plerixafor or G-CSF. P5, who was the only subject who was mavorixafor-experienced, stated a preference for Omnipod-infused plerixafor, explaining that she had to discontinue mavorixafor because of worsened gastroesophageal mavorixafor reflux and declining renal function. Moreover, she found the need to dose mavorixafor at a fixed time relative to food consumption to be a disadvantage.

## Discussion

This study demonstrates that continuous subcutaneous low dose plerixafor infusion is well-tolerated and effective at durably correcting panleukopenia in WHIM patients. The Omnipod solves several problems shared by plerixafor and insulin, including frequent dosing due to short half-lives, high cost from compounding sterile syringes, inconvenience and potential for infection from frequent injections. Compliance was excellent, and all patients preferred Omnipod over daily injections. Patient P5, who had also experienced mavorixafor, preferred Omnipod delivery of plerixafor owing to fewer side effects and no conflicts with meals.

Interestingly, we observed variable mobilization efficiency for the same leukocyte subtype among different patients and among different leukocyte subtypes for the same patient. Only P1 and P5 normalized both ANC and ALC within the prespecified low-dose range. P2 had an anomalously high baseline ALC hypersensitive to further increase by plerixafor, so that lymphocytosis accompanied ANC normalization. Conversely, for P3, P4 and P6, ANC was less responsive than ALC. Pharmacodynamic heterogeneity among subsets for mobilization might result from differences in cell type mass, cell type-specific reservoir capacity (e.g. from splenectomy in P2 and P3), CXCR4 signaling variability, pharmacokinetics, and other factors. Unexpectedly, the proportions of circulating patient neutrophils at baseline expressing CXCR1 and CXCR2 were reduced in patients, which may reflect distorted distribution dynamics, and might also contribute to neutropenia-dependent immunodeficiency. In this regard, plerixafor increased the proportion of circulating CXCR1^+^ and CXCR2^+^ neutrophils in most patients.

Since relationships among absolute leukocyte counts and pathogen susceptibility are undefined in WHIM patients, and since infections controlled by either innate or adaptive immunity may occur, which absolute cell count to choose as biomarker to optimize plerixafor dosage is also undefined. Nevertheless, continuously infused plerixafor provides flexibility over syringes and oral antagonists to finely tune dose. Whether CXCR4 antagonists provide benefits to immune responsiveness and host defense besides those postulated to accrue from redistributing mature leukocytes to blood is unknown.

Limitations of our study include the open-label design, a single center, few patients, and narrow and low dose range. Moreover, clinical efficacy is descriptive. Nevertheless, most infections were minor, and the infection rate was lower than those observed during either G-CSF or twice daily plerixafor treatment in Phase 3 (2.1 vs 3.89 and 2.84/patient-year, respectively). Moreover, all four patients with skin infections improved on drug, including all three with evaluable warts. Additional studies of HPV types, drug dose and treatment duration are needed to investigate the heterogeneity of wart responsiveness that we observed in this and our phase 3 study. In conclusion, a continuous infusion pump may be a convenient, safe, patient-preferred and potentially cost-effective means of delivering plerixafor chronically to correct panleukopenia and to improve chronic skin conditions in WHIM patients. Further assessment of safety and clinical efficacy will require larger trials, including direct comparisons to placebo, G-CSF and mavorixafor.

## Data Availability

All data produced in the present work are contained in the manuscript

## Acknowledgments

The authors thank the participants and their families for participating in this research as well as the guidance of the Independent Safety Monitoring Committee which consisted of Drs. Brian L. Kelsall, Joshua M. Farber and Michail S. Lionakis. Glenn Nardone performed the mass spectroscopy analysis of plerixafor in Omnipods. This project was supported by the Division of Intramural Research, NIAID, NIH. The content of this publication does not necessarily reflect the views or policies of the Department of Health and Human Services, and neither does the mention of trade names, commercial products, or organizations imply endorsement by the US Government. The study was also supported by Sanofi-Genzyme, Inc., which provided plerixafor.

## Authorship Contributions

D.H.M. and P.M.M. conceived of and designed the study. D.H.M., D.V., E.C., M.G.L., S.L.S., L.A.C-S. and P.M.M. provided patient care. D.F. developed the statistical analysis plan. S.M. performed flow cytometry of patient cells. D.H.M., D.F., M.C.G., S.M. and P.M.M. analyzed the data. J-L.G., Z.L. and S.M. performed the preclinical experiments of AMD3100 in mice. D.H.M. and P.M.M. wrote the first draft of the manuscript, which was supplemented by all authors.

## References

1. Tassone L, Notarangelo LD, Bonomi V, et al. Clinical and genetic diagnosis of warts, hypogammaglobulinemia, infections, and myelokathexis syndrome in 10 patients. J Allergy Clin Immunol. 2009;123(5):1170–1173, 1173 e1171-1173.

2. Beaussant Cohen S, Fenneteau O, Plouvier E, et al. Description and outcome of a cohort of 8 patients with WHIM syndrome from the French Severe Chronic Neutropenia Registry. Orphanet J Rare Dis. 2012;7:71.

3. Dotta L, Notarangelo LD, Moratto D, et al. Long-Term Outcome of WHIM Syndrome in 18 Patients: High Risk of Lung Disease and HPV-Related Malignancies. J Allergy Clin Immunol Pract. 2019;7(5):1568–1577.

4. Heusinkveld LE, Majumdar S, Gao JL, McDermott DH, Murphy PM. WHIM Syndrome: from Pathogenesis Towards Personalized Medicine and Cure. J Clin Immunol. 2019;39(6):532-556.

5. McDermott DH, Murphy PM. WHIM syndrome: Immunopathogenesis, treatment and cure strategies. Immunol Rev. 2019;287(1):91–102.

6. Dale DC, Dick E, Kelley M, Makaryan V, Connelly J, Bolyard AA. Family studies of warts, hypogammaglobulinemia, immunodeficiency, myelokathexis syndrome. Curr Opin Hematol. 2020;27(1):11–17.

7. Geier CB, Ellison M, Cruz R, et al. Disease Progression of WHIM Syndrome in an International Cohort of 66 Pediatric and Adult Patients. J Clin Immunol. 2022;42(8):1748–1765.

8. Garcia-Carmona Y, Chavez JL, Gernez Y, Geyer JT, Bussel JB, Cunningham-Rundles CMDP. Unexpected Diagnosis of WHIM syndrome in Refractory Autoimmune Cytopenia. Blood Adv. 2024.

9. Hernandez PA, Gorlin RJ, Lukens JN, et al. Mutations in the chemokine receptor gene CXCR4 are associated with WHIM syndrome, a combined immunodeficiency disease. Nat Genet. 2003;34(1):70–74.

10. Balabanian K, Lagane B, Pablos JL, et al. WHIM syndromes with different genetic anomalies are accounted for by impaired CXCR4 desensitization to CXCL12. Blood. 2005;105(6):2449–2457.

11. McDermott DH, Lopez J, Deng F, et al. AMD3100 is a potent antagonist at CXCR4(R334X), a hyperfunctional mutant chemokine receptor and cause of WHIM syndrome. J Cell Mol Med. 2011;15(10):2071-2081.

12. Mc Guire PJ, Cunningham-Rundles C, Ochs H, Diaz GA. Oligoclonality, impaired class switch and B-cell memory responses in WHIM syndrome. Clin Immunol. 2010;135(3):412–421.

13. McDermott DH, Liu Q, Ulrick J, et al. The CXCR4 antagonist plerixafor corrects panleukopenia in patients with WHIM syndrome. Blood. 2011;118(18):4957–4962.

14. Laberko A, Deordieva E, Krivan G, et al. Multicenter Experience of Hematopoietic Stem Cell Transplantation in WHIM Syndrome. J Clin Immunol. 2022;42(1):171–182.

15. Rosenberg PS, Alter BP, Bolyard AA, et al. The incidence of leukemia and mortality from sepsis in patients with severe congenital neutropenia receiving long-term G-CSF therapy. Blood. 2006;107(12):4628–4635.

16. McDermott DH, Pastrana DV, Calvo KR, et al. Plerixafor for the Treatment of WHIM Syndrome. N Engl J Med. 2019;380(2):163–170.

17. McDermott DH, Velez D, Cho E, et al. A phase III randomized crossover trial of plerixafor versus G-CSF for treatment of WHIM syndrome. J Clin Invest. 2023;133(19).

18. Badolato R, Alsina L, Azar A, et al. A phase 3 randomized trial of mavorixafor, a CXCR4 antagonist, for WHIM syndrome. Blood. 2024;144(1):35–45.

19. McDermott DH, Liu Q, Velez D, et al. A phase 1 clinical trial of long-term, low-dose treatment of WHIM syndrome with the CXCR4 antagonist plerixafor. Blood. 2014;123(15):2308–2316.

20. Winer KK, Zhang B, Shrader JA, et al. Synthetic human parathyroid hormone 1-34 replacement therapy: a randomized crossover trial comparing pump versus injections in the treatment of chronic hypoparathyroidism. J Clin Endocrinol Metab. 2012;97(2):391–399.

21. Balabanian K, Brotin E, Biajoux V, et al. Proper desensitization of CXCR4 is required for lymphocyte development and peripheral compartmentalization in mice. Blood. 2012;119(24):5722–5730.

22. Gao JL, Yim E, Siwicki M, et al. Cxcr4-haploinsufficient bone marrow transplantation corrects leukopenia in an unconditioned WHIM syndrome model. J Clin Invest. 2018;128(8):3312–3318.

23. Majumdar S, Pontejo SM, Jaiswal H, et al. Severe CD8+ T Lymphopenia in WHIM Syndrome Caused by Selective Sequestration in Primary Immune Organs. J Immunol. 2023;210(12):1913–1924.

24. Rosenkilde MM, Gerlach LO, Jakobsen JS, Skerlj RT, Bridger GJ, Schwartz TW. Molecular mechanism of AMD3100 antagonism in the CXCR4 receptor: transfer of binding site to the CXCR3 receptor. J Biol Chem. 2004;279(4):3033–3041.

