## Supplemental Appendix for "Continuous Infusion of the CXCR4 Antagonist Plerixafor for WHIM Syndrome"

McDermott et al.

**Methods**

**Stability testing of plerixafor in the Omnipod delivery system**

The analysis was done on an Agilent 1260 analytical HPLC system (Santa Clara, CA) using a Luna C_18_, 3-micron, 4.6 x 200 mm column. Equilibration was done in 90% 0.1% trifluoroacetic acid - 10% acetonitrile at 0.5 ml/min. The column was developed with a gradient of acetonitrile from 10% to 54% over 13 minutes after a 5-minute wash. The injection volumes were 5 μl and peak detection and integration was done at 220 nm. Five-point calibration curves from 1 to 4 μg plerixafor were linear. Plerixafor from the infusion pump or from controls was diluted 50X in either 6M urea or 0.1% TFA for the analysis with nearly identical results. Sample concentrations were determined with 4 replicate dilutions. Intact mass analysis of plerixafor was done with a 4000 Q Trap mass spectrometer (SCIEX Corporation, Framingham, MA) at 3500 V in positive mode by direct infusion through an in-line Optimize small molecule micro-trap at 20 μl/min. The spray solvent was 20% 5 mM ammonium acetate, pH 4.5 and 80% acetonitrile. Scanning was done from 100 to 530 m/z over 342 cycles. Under these conditions the +1 and +2 ions were dominant at 503.5 and 252.2 respectively. Plerixafor was placed in a Pod that was held at 37°C for 3 days in an incubator and then tested for concentration (biocompatibility) and breakdown products (thermal stability) versus the reference product provided by the manufacturer.

**Flow cytometry analysis of chemokine receptor expression on circulating neutrophils**

Analysis was performed on freshly drawn peripheral blood samples from the WHIM patients in this study and 33 adult healthy donors recruited at the NIH-CC. Sampling timepoints were day 0 before administering plerixafor, 3-6 days and ~6 months after starting plerixafor, and at the end of the study. First, 100-200 µL of whole blood was stained with Zombie UV (BioLegend, San Diego, California) or LIVE/DEAD™ Fixable Aqua Dead Cell Stain (Thermo Fisher Scientific, Waltham, MA) for 10 min at room temperature. Next, Human TruStain FcX™ was added for Fc receptor blocking, followed by mAbs against human CD66b (clone: G10F5), CXCR1 (clone: 8F1/CXCR1), CXCR2 (clone: 5E8/CXCR2), CXCR4 (clone: 12G5), mouse IgG1 (clone: MOPC-21), mouse IgG2a (clone: MOPC-173) and mouse IgG2b (clone: MG2b-57) (all from BioLegend). ACK lysis buffer (Quality Biological Inc., Gaithersburg, MD) was added to lyse RBCs. Cells were washed with PBS, fixed with 4% PFA for 30 min, resuspended in PBS and analyzed with a BD Fortessa flow cytometer and FlowJo software (BD Bioscience, Franklin Lakes, NJ).

**Supplemental Tables**

**Supplemental Table 1.** Summary of study subject infection history. Severity code: 0, no infections; 1+, non-recurrent infection; 2+, recurrent infection; 3+, recurrent infection with documented evidence of end organ damage (e.g. bronchiectasis, hearing loss, tooth loss, blindness).

|  |  |  | **Prior History of Infection by Site and Severity** | | | | | | | |
| --- | --- | --- | --- | --- | --- | --- | --- | --- | --- | --- |
| **Patient** | **Prior**  **G-CSF Rx (yrs)** | **Prior Ig Rx** | **Sinus** | **Middle Ear** | **Lung** | **Skin** | **GU** | **Blood** | **Teeth** | **Comment/Other** |
| **P1** | 30 | 36 | 3+ | 3+ | 3+ | 1+ | 1+ | 0 | 2+ | *P. aeruginosa* colonization of airway |
| **P2** | 1 | 14 | 2+ | 3+ | 3+ | 3+ | 1+ | 1+ | 1+ | Osteomyelitis, Septic arthritis, *Aspergillus sp.* |
| **P3** | 21 | 8 | 2+ | 2+ | 2+ | 3+ | 0 | 2+ | 0 | Pneumococcal sepsis, perirectal abscess |
| **P4** | 12 | 0 | 2+ | 2+ | 3+ | 0 | 0 | 1+ | 0 | *Toxoplasma gondii* chorioretinitis,  endocarditis |
| **P5** | episodic | 0 | 2+ | 3+ | 3+ | 1+ | 2+ | 1+ | 3+ | Sepsis |
| **P6** | 14 | 14 | 2+ | 3+ | 3+ | 2+ | 0 | 1+ | 2+ | Septic arthritis,  Sepsis, Tinea |

Abbreviations: GU, genitourinary tract; Rx, treatment; yrs, years; Ig, immunoglobulin supplementation.

**Supplemental Table 2.** Characterization of wart areas on WHIM patients treated with continuous

plerixafor infusion. 1+, a single wart; 2+, a few warts in a group; 3+, a large wart area; 4+, a wart

area extensively covering an entire body part; PR, partial response (>=50% reduction in size); CR, complete response; ne, not

evaluable; NR, no response; L, left; R, right; F, finger; DIP, distal interphalangeal joint; PIP,

proximal interphalangeal joint.

| **Patient** | **Location** | **Type** | **Burden** | **Response** | **Additional Treatment** |
| --- | --- | --- | --- | --- | --- |
| **P1** | R 5^th^ F subungual | Verruca vulgaris | 2+ | PR | none |
|  | R 2^nd^ F DIP & PIP joints | Verruca vulgaris | 2+ | PR | none |
|  | R 2^nd^ MCP joint | Verruca vulgaris | 2+ | PR | none |
|  | R palm | Flat papule | 1+ | PR | none |
|  | R wrist | Verruca vulgaris | 3+ | PR | none |
|  | L 5^th^ F subungual | Verruca vulgaris | 2+ | PR | none |
|  | L 2^nd^ F DIP joint | Verruca vulgaris | 1+ | ne | none |
|  | R upper lip | Flat papule | 1+ | CR | none |
| **P2** | R wrist | Clusters of verruca plana | 3+ | PR | none |
|  | L wrist | Clusters of verruca plana | 2+ | CR | none |
|  | R dorsal hand | Groups of verruca plana | ne | ne | Imiquimod, month 8-9 |
|  | L dorsal hand | Groups of verruca plana | ne | ne | none |
|  | Proximal R 3^rd^ dorsal F | Groups of verruca plana | 3+ | PR | none |
|  | L anterior thigh | verruca plana | 1+ | PR | none |
|  | R pretibial area | Verrucous plaque | 1+ | PR | none |
|  | Genitalia | Condyloma accuminata | 2+ | NR | Imiquimod, month 8-9 |
| **P5** | Dorsum of R hand and fingers | Verrucous papules | 4+ | NR | Imiquimod, 3 times/week,  months 0-2 and 6-10 |
|  | Both anterior thighs to anterior lower legs | EV-like verruca | 4+ | PR | none |
| **P6**^a^ | R dorsal hand and fingers | Flat papules | 3+ | ne | none |
|  | L dorsal hand and fingers | Flat papules | 3+ | ne | none |
|  | R volar wrist | Flat papules | 2+ | ne | none |
|  | R proximal medial thigh to inguinal area | Pedunculated papules | 3+ | ne | none |
|  | Perianal area | Pedunculated papules | 3+ | ne | none |

^a^Patient P6 withdrew from the study after 2 months for social reasons.

**Supplementary Figure 1.** Timeline for quantification of chemokine receptor expression on blood leukocytes of WHIM patients. Leukocytes in freshly obtained blood samples were stained for chemokine receptor expression for the patients listed on the left at the adjacent timepoints indicated by the arrows. G, on G-CSF treatment; Baseline, day 0; P, 3-6 days and ~6 and ~12 months after starting continuous plerixafor administration. N. B. Patient P6 dropped out for social reasons after 2 months of plerixafor treatment, and the 12 month measurement was not done for patients P4 and P5.

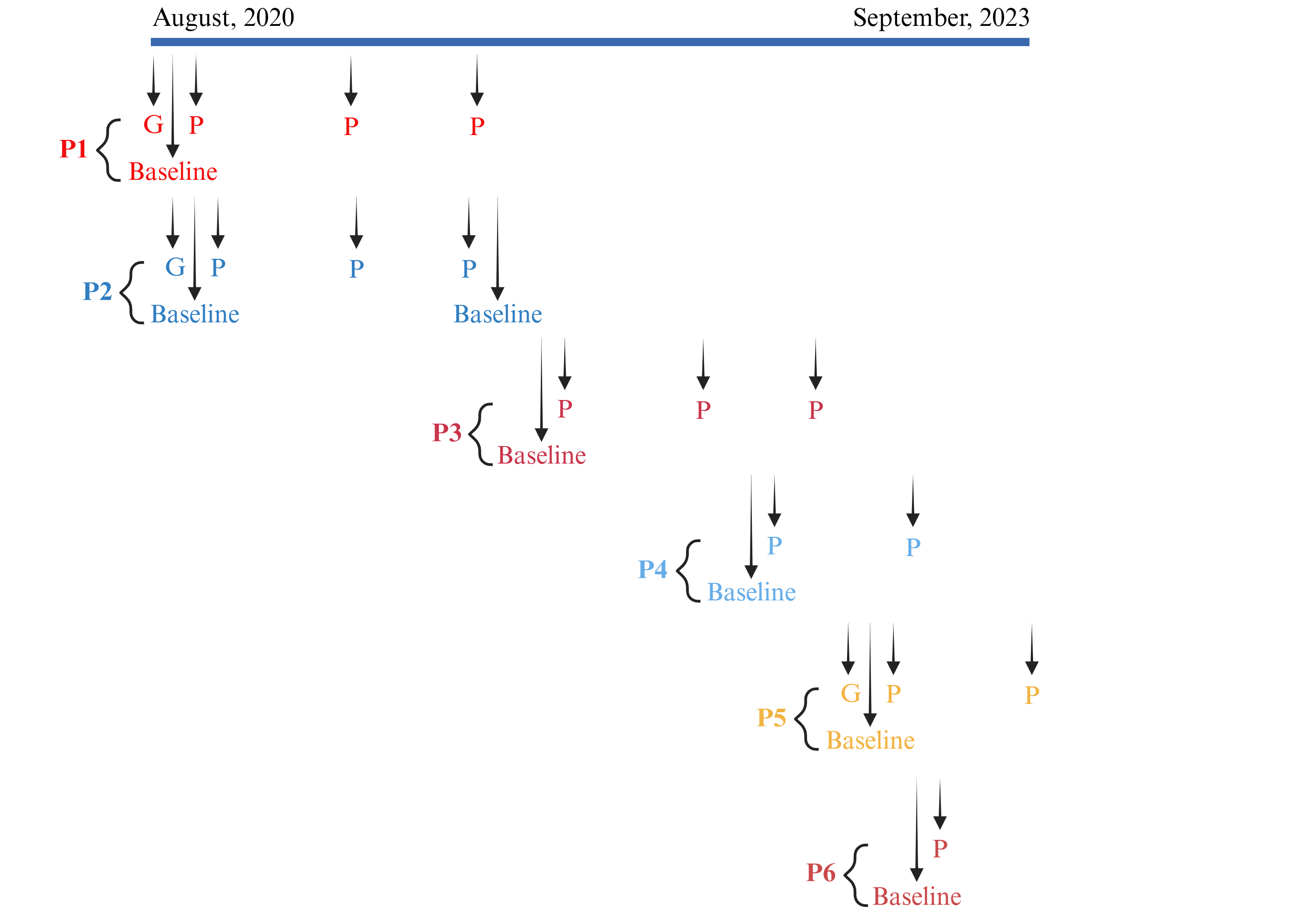

**Supplemental Figure 2.** Gating strategy for quantification of chemokine receptor expression on circulating neutrophils in WHIM patients and healthy donors.

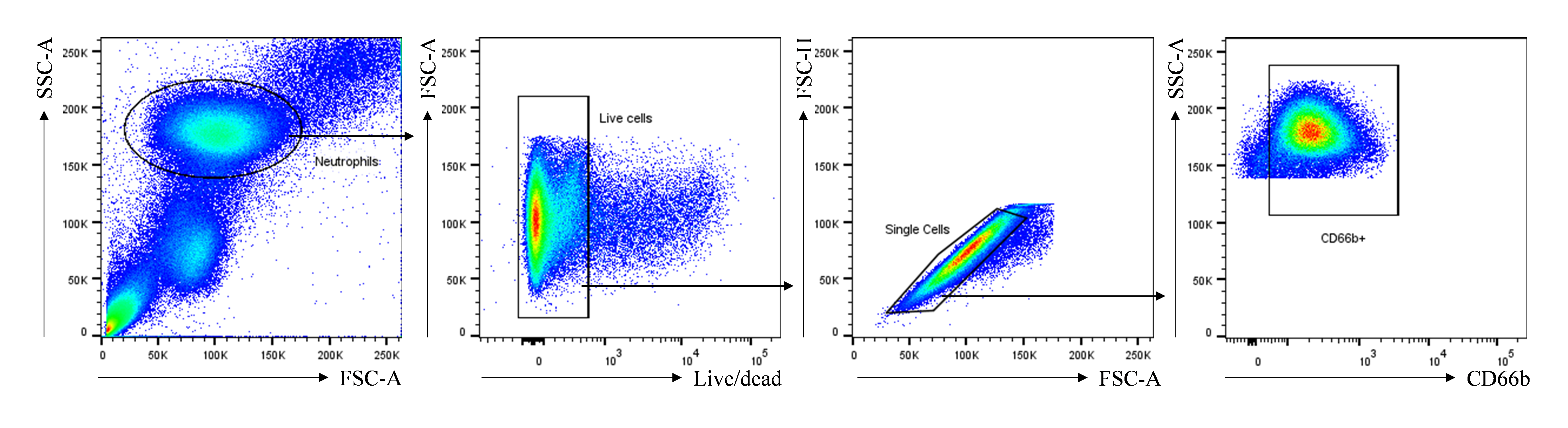

National Institute of Allergy and Infectious Diseases (NIAID)/National Institutes of Health (NIH)

Laboratory of Molecular Immunology (LMI)

Protocol Face Sheet:

Principal Investigator (PI):	David H. McDermott, MD

Institute/Branch:	NIAID/LMI

Address:	Bldg 10, Room 11N107, MSC 1886, 10 Center Drive, Bethesda, MD 20892

Protocol Number:	09-I-0200

Version Description:	20

Version Date	Aug 28, 2024

Protocol Title:	A Study of Mozobil^TM^ in the Treatment of Patients with WHIMS

Abbreviated Title:	WHIMS and Mozobil^TM^

Institutional Review Board (IRB) Name:	NIH

Institute Name:	NIH

Proposed Dates:	Start-6/1/2009	End 6/30/2028

Total Subjects to be accrued:	20

Ionizing Radiation Use:	Medically indicated

Is tissue being collected for Research Purposes?	Yes (optional wart/bone marrow biopsies)

Location of the Study:	NIH Clinical Research Center

IND:	Exempted

Name of Agent/Device:	Mozobil^TM^

Sponsor:	none

Holder:	none

**Confidentiality Statement**

This document is confidential and is to be distributed for review only to investigators, potential investigators, consultants, study staff, and applicable independent ethics committees or institutional review boards. The contents of this document shall not be disclosed to others without written authorization from NIAID (or others, as applicable), unless it is necessary to obtain informed consent from potential study participants.

Table of Contents

Protocol Face Sheet:	1

Table of Contents	2

List of Abbreviations	5

Précis	7

Statement of Compliance	7

1	INTRODUCTION	8

1.1	Study Overview	8

1.2	Study Objectives	8

1.3	Hypothesis	8

1.4	Background	8

1.4.1	Pathogenesis and Etiology	8

1.4.2	Description of the Study Agent	10

1.4.3	Clinical Pharmacology/Mechanism of Action	10

1.4.4	Pharmacokinetics	11

1.4.5	Formulation, Packaging and Labeling	11

1.4.6	Summary of Previous Pre-Clinical Studies	11

1.4.7	Summary of Relevant Clinical Studies	11

1.4.8	Adverse Reactions	12

1.4.9	Warnings and Precautions	12

1.4.10	Drug Interactions	13

1.5	Scientific and Clinical Justification	13

1.6	Qualifications of Investigators	13

1.7	Conflict of Interest	13

1.8	Conduct of the Protocol	13

1.9	Observed Risks and Incidents in an NIH Study with Plerixafor and G-CSF	14

2	STUDY DESIGN	14

2.1	Overview	14

2.1.1	Phase A, B, or A & B: Dose Escalation (optional)	14

2.1.2	Phase C: Chronic Dosing with Syringes (optional)	15

2.1.3	Phase D: Trial Using an Infusion Pump – in place of syringes (optional)	15

2.2	Study Endpoints	16

2.2.1	Primary Endpoints	16

2.2.2	Exploratory Endpoints	16

2.3	Sample Size Justification	16

3	SUBJECT ENROLLMENT	16

3.1	Patient Recruitment	16

3.2	Inclusion/Eligibility Criteria	17

3.3	Exclusion Criteria	17

3.4	Study Enrollment Procedures	17

3.4.1	Informed Consent	17

3.4.2	Justification for Exclusion of Women, Minorities, and Children (Special Populations)	19

3.4.3	Request for Waiver of Informed Consent for Secondary Research from NIH IRB Protocol 14-I-0185	19

4	STUDY IMPLEMENTATION	19

4.1	Criteria for Withdrawal	19

4.1.1	Voluntary Withdrawal	19

4.1.2	Involuntary Withdrawal	19

4.2	Study Parameters	20

4.2.1	Pretreatment Evaluations Within 30 Days	20

4.2.2	Active Phase A (Dose Escalation Phase Inpatient)	21

4.2.3	Active Phase B (Response to Maximum Dose - Inpatient)	22

4.2.4	Chronic Phase with Syringes (Phase C)	23

4.2.5	Pilot Trial with Infusion Pump (Phase D)	24

4.2.6	Phase C and D Safety Monitoring	24

4.2.7	Study Completion Visit	24

4.3	Collaboration with Other Investigators	25

4.4	Failure to Respond to Mozobil^TM^	25

4.5	Dose/Schedule Modifications for a Participant	25

4.6	Study Drug Acquisition	26

4.7	Prohibited Medications and Procedures	26

4.8	Subject Monitoring and Supportive Care	26

4.9	Temporary Suspension Rules for the Protocol	27

4.10	Stopping Rules for an Individual Participant/Cohort	27

5	ANALYSIS OF STUDY	27

6	POTENTIAL HAZARDS AND DISCOMFORTS	29

6.1	Potential Risks of Mozobil^TM^	29

6.2	Risks and Discomforts of Phlebotomy	30

6.3	Risks and Discomforts of Wart Biopsy	30

6.4	Risks and Discomforts of Bone Marrow Biopsy	30

6.5	Risks and Discomforts of HIV Testing	31

6.6	Risks of Stopping G-CSF/GM-CSF	31

6.7	Risk of Genetic Testing	31

6.8	Risks and Discomforts of the Infusion Pump Device	31

7	STUDY POTENTIAL BENEFIT	32

8	HUMAN SUBJECT PROTECTIONS	32

8.1	Institutional Review Board	32

8.2	Protocol Amendments and Study Termination	32

8.3	Privacy and Confidentiality Provisions	33

8.4	Considerations of Alternatives to Participation	33

8.5	Policy Regarding Research-Related Injuries	33

8.6	Subject Remuneration	33

9	SAFETY ASSESSMENT	34

9.1	Reporting Procedures	34

9.2	Reporting of Pregnancy	34

9.3	Toxicity Criteria	34

10	DATA AND SAFETY MONITORING	35

11	PROTOCOL MONITORING PLAN	35

12	PLAN FOR USE AND STORAGE OF BIOLOGICAL SAMPLES	35

12.1	Use of Stored Samples and Data	35

12.2	Disposition of Stored Samples and Data	35

13	PUBLICATION POLICY	36

14	REFERENCES	36

APPENDIX A: TOXICITY TABLE	39

APPENDIX B: SCHEDULE OF PROCEDURES/EVALUATIONS for Phase A and/or B	42

APPENDIX C: DOSING OF MOZOBIL^TM^ IF THERE IS RENAL IMPAIRMENT	43

APPENDIX D: FDA APPROVED PACKAGE INSERT FOR MOZOBIL^TM^	44

APPENDIX E: NIAID/LMI SAFETY MONITORING COMMITTEE	45

APPENDIX F: SCHEDULE OF EVENTS FOR PHASES C AND D	47

APPENDIX G: A SAMPLE OF PLERIXAFOR DOSES, INFUSION RATES, AND STEADY STATE CONCENTRATIONS	48

APPENDIX H: PLERIXAFOR INFUSION	49

List of Abbreviations

| AI | Associate Investigator |
| --- | --- |
| ALC | Absolute lymphocyte count |
| ANC | Absolute neutrophil count |
| CFR | Code of Federal Regulations |
| CBC | complete blood count with differential |
| CD | chronic dose. The dose of Mozobil to be given twice / day in Phase C. Defined as ½ the minimally effective dose |
| CrCl | creatinine clearance (calculated) |
| CRIS | Clinical Research Information System (for NIH Clinical Center) |
| CSO | Clinical Safety Office |
| CXCL12 | CXC chemokine ligand 12, aka SDF-1 |
| CXCR4 | CXC chemokine receptor 4 |
| ECHO | Echocardiogram |
| EKG | Electrocardiogram |
| FDA | Food and Drug Administration |
| FWA | Federal Wide Assurance |
| GCP | Good Clinical Practices |
| G-CSF | Granulocyte-colony stimulating factor |
| GM-CSF | Granulocyte macrophage-colony stimulating factor |
| HIV | Human Immunodeficiency Virus |
| HPV | Human papillomavirus |
| HRPP | Human Research Protection Program |
| HSC | Hematopoietic Stem Cell |
| ICH | International Council on Harmonisation of Technical Requirements for Pharmaceuticals for Human Use |
| IgA, IgE, IgG, IgM | immunoglobulins A, E, G, M |
| IND | Investigational New Drug |
| IRB | Institutional Review Board |
| IVIG | intravenous immunoglobulin |
| MED | Minimally effective dose. The dose of Mozobil in phase A that causes a 2-fold rise in ANC AND ANC>500 |
| MM | multiple myeloma |
| MOP | Manual of Procedures/Manual of Operations |
| NCI | National Cancer Institute, NIH |
| NHL | non-Hodgkin’s lymphoma |
| NIAID | National Institute for Allergy and Infectious Diseases, NIH |
| NIH | National Institutes of Health |
| OHRP | Office for Human Research Protection |
| PI | Principal Investigator |
| PT | prothrombin time |
| PTT | partial thromboplastin time |
| RCHSP | Regulatory Compliance and Human Subjects Program |
| SCN | Severe Congenital Neutropenia |
| SDF-1 | Stromal cell-derived factor 1, aka CXCL12 |
| SQ | subcutaneous |
| WBC | white blood cell count |
| WHIMS | Warts, Hypogammaglobulinemia, Infections, and Myelokathexis Syndrome. |
| UP | Unanticipated Problem |
| UPnonAE | Unanticipated Problem that is not an Adverse Event |

Précis

Mozobil™ (plerixafor injection, Genzyme/Sanofi) is a Food and Drug Administration–approved medication to mobilize CD34^+^ hematopoietic stem cells prior to apheresis and use in autologous transplantation in non-Hodgkin lymphoma and multiple myeloma when used in conjunction with granulocyte-colony stimulating factor (G-CSF). The drug’s mechanism of action is the specific and reversible inhibition of the chemokine receptor, CXCR4, expressed on CD34^+^ cells and other leukocytes. This inhibition interferes with the binding of stromal cell derived factor-1 (SDF-1), which is constitutively expressed on bone marrow stromal cells and appears to cause direct and indirect cellular adhesive interactions. Severe congenital neutropenia is a rare inherited disorder in which the affected individuals develop chronic or cyclical neutropenia with circulating counts below 500 cells/microliter blood. This disorder may result from a variety of genetic defects in progenitor- or neutrophil-expressed genes such as elastase, *CXCR4*, *G6PC3*, etc. Myelokathexis is the abnormal retention of mature leukocytes in the bone marrow and is seen in some types of severe chronic neutropenia such as warts, hypogammaglobulinemia, infections, and myelokathexis syndrome (WHIMS). WHIMS is a rare primary immunodeficiency, which is known to be caused by mutations that enhance CXCR4 signaling. Our hypothesis is that Mozobil™ can be used safely to partially block CXCR4 and treat neutropenia resulting from myelokathexis at doses considerably lower than that being used for CD34^+^ cell mobilization. This new treatment could also improve other aspects of the disease such as frequent infections, warts, and hypogammaglobulinemia. To test this hypothesis, we propose this trial of Mozobil™ in adults with WHIMS, examining safety and absolute neutrophil count as the primary endpoint. Mozobil™ is injected subcutaneously and will be injected via syringes (up to 84 months) or via an infusion pump (pilot trial of up to 10 subjects for a 24‑month period).

Statement of Compliance

The study will be conducted in accordance with the design and specific provisions of this IRB‑approved protocol, in accordance with the ethical principles that have their origin in the Declaration of Helsinki, and that are consistent with Good Clinical Practices (GCP) and the applicable regulatory requirement(s). The PI will assure that no deviation from or changes to the protocol will take place without prior documented approval from the IRB, except where necessary to eliminate an immediate hazard(s) to the trial participants. The PI will promptly report to the IRB any changes in research activity and all unanticipated problems involving risk to human subjects, or others.

### INTRODUCTION

#### Study Overview

This is an open-label, dose ranging study of the safety and efficacy of Mozobil™ in the treatment of adults with WHIMS that consists of 4 different phases that can be completed consecutively or separately. After a wash-out period to remove all effects of granulocyte colony stimulating factor (G-CSF) or granulocyte macrophage colony stimulating factor (GM-CSF), in Phase A we will administer Mozobil™ at increasingly higher doses (dose escalation) until the neutrophil count is normalized or until the maximum Food and Drug Administration (FDA)-approved dose is reached (0.24 mg/kg/day [or 0.16 mg/kg/day if participant has renal impairment]). Our target value is an absolute neutrophil count (ANC) of ~4000 cells/microliter of blood, which is the mean for children and adults. Complete blood count with differential (CBC) will be performed frequently and the dose increased until that target is achieved or until the current FDA-approved dose is reached. In Phase B, either the dose that normalized neutrophil count or the current FDA-approved dose will be administered, and measurements of the leukocyte response and/or drug levels will be performed. Participants may also participate in a chronic treatment phase of the study where they receive a minimally effective dose (MED) of Mozobil^TM^ for up to 84 months via syringes, or in a pilot trial with the drug delivered via a disposable infusion pump for up to 24 months.

There will be no IND obtained for the use of any of the commercial agents used in this study. This study meets the criteria for exemption for an IND as this investigation is not intended to support a new indication for use or any other significant change to the labeling; the drugs are already approved and marketed and the investigation is not intended to support a significant change in advertising; and the investigation does not involve a route of administration or dosage level in use in a patient population or other factor that significantly increases the risks (or decreases the acceptability of the risks) associated with the use of the drug product.

#### Study Objectives

The objectives of this study are to assess the safety and efficacy of Mozobil™ in the treatment of neutropenia resulting from the abnormal retention of mature neutrophils and other white blood cells in the bone marrow (myelokathexis).

#### Hypothesis

Mozobil™ treatment can be given safely and results in a clinically significant increase in ANC in patients with neutropenia due to myelokathexis.

#### Background

##### Pathogenesis and Etiology

Neutropenia is a reduction in a particular type of circulating white blood cell called a neutrophil that is a critical part of host defense against infection. Reductions below 1000 cells/microliter of blood are known to raise the risk of infection. Prolonged reductions below 500 cells/microliter of blood are particularly dangerous and greatly increase the susceptibility to bacterial and fungal infections. Severe congenital neutropenia (SCN) is a rare but serious inherited immunodeficiency caused by mutations in a variety of genes important in the development, lifespan, and release of neutrophils.[1, 2] One of these genes, CXCR4, is a chemokine receptor with important roles in immune system development and the adherence and homing of human CD34^+^ hematopoietic stem cells (HSCs) and other leukocytes to the bone marrow stroma.[3] Excessive functioning of this gene is one of the causes of SCN that we seek to treat in this study.

WHIMS is a rare congenital immunodeficiency caused by various mutations that increase the signaling of CXCR4.[4, 5] Most of the genetically characterized cases to date have involved truncation mutations in the carboxyl terminus of the receptor that result in abnormal desensitization and are inherited in an autosomal dominant manner.[6, 7] One individual has been found to have abnormal expression of another gene, *GRK3*, involved in CXCR4 desensitization.[8] Thus, WHIMS can be thought of as a disease caused by excessive CXCR4 function. It is possible that other genetic mutations may also cause a similar syndrome.

The disease manifests with multiple signs that include warts, hypogammaglobulinemia, infections, and myelokathexis. The warts are difficult to treat and eventually cause serious and sometimes fatal cancers. The hypogammaglobulinemia is moderate and variable, but patients have been reported to have poor immunologic memory after vaccination, and the resulting low levels of immunoglobulins predispose to frequent sinopulmonary infections. The primary clinical finding is typically a severe and constant neutropenia due to the abnormal retention of mature neutrophils in the bone marrow. This further predisposes to typical bacterial skin and soft tissue and sinopulmonary infections. During these infections, the neutropenia often improves and may temporarily normalize, suggesting that the inhibition of marrow release can be overcome in the presence of infection. However, the end result of these frequent infections can be very serious, with loss of hearing, teeth, and lung function. Older patients can develop severe bronchiectasis resembling cystic fibrosis, and respiratory failure. Case reports of fatal meningitis and Epstein‑Barr virus–induced lymphoma in patients with WHIMS also indicate other significant immunological problems.[9, 10]

Current therapeutic recommendations are based on limited clinical experience and no controlled clinical trials in WHIMS patients, but involve daily or every other day injection of G-CSF to normalize neutrophil counts, and monthly infusion of intravenous immunoglobulin (IVIG).[7] These therapies are expensive, non-specific, have significant side effects and toxicities, and do not fully correct the patient’s problems, especially warts and human papillomavirus (HPV)‑related cancers.

Two recent short-term Phase I dose escalation studies of Mozobil^TM^ in a total of 9 patients, one conducted by our group using a previous version of this protocol, demonstrated that the drug could safely mobilize not only neutrophils, but also all other leukocyte subsets that are decreased in the blood of WHIMS patients.[11, 12] Three WHIMS patients that received Mozobil^TM^ 0.02‑0.04 mg/kg/day given as a twice daily subcutaneous (SQ) injection for 6 months showed that these hematopoietic effects were durable. Treatment was associated with only 2 infections in one patient, and appeared to reduce wart burden, with no adverse events (AEs).[13] This finding has been confirmed in an additional 3 patients treated for 19 to 52 months with plerixafor using this protocol.[14]. In addition, we have completed a randomized masked trial (NIH IRB protocol 14-I-0185, “A Phase III, Double-blind, Randomized, Crossover Study of Plerixafor Versus G‑CSF in the Treatment of Patients with WHIM Syndrome”) comparing Neupogen (filgrastim or G-CSF) versus Mozobil (plerixafor) in 19 participants with WHIM syndrome. In this crossover trial, participants were randomized to one drug or the other and treated for 14 months; following a brief washout period, participants were then treated with the second study drug for another 14 months. Both study drugs were given as a twice‑daily SQ injection.[15] This was a monitored trial with children (aged 10 years and above) and adults, and was closed in 2023. We found that plerixafor was superior to G-CSF for maintaining lymphocytes >1000 cells/microliter and non-inferior for keeping neutrophils >500, while the infection rate and severity was nearly identical when comparing plerixafor to the standard of care for SCN.

In the above studies, lymphocyte mobilization was robust but rapidly peaked (3-6 hours) after injection and then fell back to baseline (9-12 hours). Based on unpublished observations in a WHIMS murine model (Majumdar, et al.), there is reason to believe that continuous administration of plerixafor using a SQ infusion pump may be superior to twice daily injection. We will repurpose a disposable, sterile, self-contained unit (OmniPod®) to deliver plerixafor as has been done in a prior successful study at NIH for parathyroid hormone.[16] This device has been FDA-approved to continuously deliver several types of U-100 insulin since 2005 and a delayed single dose of a long-acting G-CSF drug (pegfilgrastim) as Neulasta OnPro® since 2015 and has an excellent safety record with over 100,000 current users.[17] Each device will be filled aseptically with the contents of a single plerixafor vial and used for no more than 3 days before disposal.

##### Description of the Study Agent

Mozobil^TM^ (plerixafor injection, previously known as AMD3100, JM3100, and SDZ SID791) has been approved by the FDA for use in combination with G-CSF to mobilize HSCs to the peripheral blood for collection and subsequent autologous transplantation in patients with non‑Hodgkin’s lymphoma (NHL) and multiple myeloma (MM). Mozobil™ was initially developed as an antiviral compound for the treatment of human immunodeficiency virus (HIV).[18] It was found to be a specific and reversible inhibitor of the chemokine receptor and HIV coreceptor CXCR4, and its plasma concentrations closely correlated with the ability to inhibit X4 HIV viral entry into target cells.[19] While initial studies showed that it was a safe and effective antiviral drug in animals and humans, CXCR4-using virus is not common in initial HIV infection and the drug is not orally bioavailable, limiting its usefulness for treating HIV. However, in early human studies, it was noted that the drug caused a dramatic and rapid increase in white blood cell count (WBC).

This was consistent with other studies in mice showing that CXCR4 is a strong bone marrow retention factor for leukocytes in general and neutrophils and CD34^+^ HSCs specifically. This prompted successful trials of Mozobil™ in animals and humans for the purpose of CD34^+^ HSC mobilization. This is a procedure in which an apheresis of the donor is done to separate and remove the CD34^+^ cells, which allow immune system reconstitution after high dose chemotherapy for cancer. Mozobil™ is effective either by itself or in combination with granulocyte-colony stimulating factor (G-CSF), which is the agent that has traditionally been used for this purpose. In Phase III clinical studies that led to its licensure in the US, it was found that there was a dramatic, statistically significant increase in the percentage of difficult to mobilize (because of prior chemotherapy) individuals who reached the target numbers of cells collected when Mozobil™ was used with G-CSF rather than just G-CSF alone.

##### Clinical Pharmacology/Mechanism of Action

Mozobil™ is an inhibitor of the CXCR4 chemokine receptor and blocks binding of its cognate ligand, stromal cell-derived factor-1 (SDF-1). Both SDF-1 and CXCR4 are recognized to play a role in the trafficking and homing of human HSCs to the marrow compartment. Once in the marrow, stem cell CXCR4 can act to help anchor these cells to the marrow matrix, either directly via SDF-1 or through the induction of other adhesion molecules. Treatment with Mozobil™ resulted in leukocytosis and elevations in circulating hematopoietic progenitor cells in mice, dogs, and humans.

##### Pharmacokinetics

Pharmacokinetic studies in normal volunteers and cancer patients have shown that the drug is rapidly absorbed after SQ injection.[20, 21] Peak plasma concentration is observed at 30‑60 minutes after a single 0.24-mg dose and the half-life has been estimated at approximately 6 hours. The drug is eliminated renally, with 70% of the drug excreted unchanged in the urine in 24 hours. There are no reported drug-drug interactions, and studies to date have found no interactions with the cytochrome P450 system. The drug has been found to be teratogenic in animals and could be excreted in breast milk and is therefore contraindicated in pregnant women or nursing mothers but was negative on an in vitro test of mutagenesis. The teratogenicity is most likely a direct effect of blocking CXCR4 since animal studies have found the receptor to be critical for several aspects of embryogenesis and immune system development.[22-24]

##### Formulation, Packaging and Labeling

Mozobil^TM^ (plerixafor injection) is a sterile, preservative-free, clear, and colorless to pale yellow isotonic solution for SQ injection. Each mL of the sterile solution contains 20 mg of plerixafor. Each single-use vial is filled to deliver 1.2 mL of the sterile solution that contains 24 mg of plerixafor and 5.9 mg of sodium chloride in water for injection. Plerixafor is an HSC mobilizer with a chemical name l, 1'-[1,4- phenylenebis (methylene)]-bis-1,4,8,11-tetraazacyclotetradecane. It has the molecular formula C_28_H_54_N_8_. The molecular weight of plerixafor is 502.79 g/mol. Storage at room temperature is recommended (25 °C) with excursions from 15-30 °C acceptable with unopened and properly stored glass vials having a usable shelf‑life of at least 3 years. The chemical stability of plerixafor in room temperature or refrigerated plastic and glass sterile syringes has been also previously demonstrated for extended periods (months to years) by the NIH pharmacy and others.[25-27] We may also use a disposable infusion pump for delivery in some patients.

##### Summary of Previous Pre-Clinical Studies

In vitro studies have demonstrated that Mozobil^TM^ is specific for CXCR4 and is a reversible inhibitor of the natural ligand’s (SDF-1 also known as CXCL12) ability to bind the receptor.[19] Work at NIH in our laboratory has indicated that Mozobil™ has a very similar ability to bind and block the signaling of normal wild-type CXCR4 receptor, just as it binds to the mutant receptor that causes WHIMS. Long-term administration to mice, rats, and monkeys was found to be safe; however, in high doses (10 times highest human dose) in pregnant rats, the drug was found to be teratogenic causing fetal mortality. Also, in rats, increased spleen weight was observed after 2 to 4 weeks of daily administration at high doses (4 times the highest human dose).

##### Summary of Relevant Clinical Studies

Mozobil™ in combination with G-CSF has been used in more than 980 patients enrolled in 16 clinical studies for the purpose of stem cell mobilization as reported by the manufacturer, Genzyme/Sanofi, to the FDA. In one randomized study of 298 patients with NHL where all patients were given 0.24 mg/kg of the drug SQ daily for 1 to 7 days (median, 2 days), 59% of those who received the drug combination were able to achieve the collection goal of 5 million CD34^+^ cells/kg versus only 20% who received G-CSF alone (p<0.001). In a second randomized study, 302 patients with MM were mobilized in the same way, and 72% achieved the collection goal of 6 million CD34^+^ cells/kg versus only 34% of those who received G-CSF alone (p<0.001).

The most frequent side effects/adverse reactions were mild injection site reactions seen in 34% of patients. These injection site reactions included erythema, hematoma, hemorrhage, induration, inflammation, irritation, pain, paresthesia, pruritus, rash, swelling, and urticaria. While frequent, such reactions were generally mild and resolved without treatment in minutes. Other common adverse reactions seen in cancer patients given this drug in combination with G-CSF were mainly gastrointestinal (nausea, vomiting, diarrhea, constipation, abdominal pain/distention) but also include arthralgia, dizziness, headache, and fatigue (see Package Insert, APPENDIX D below). Mild to moderate systemic reactions were seen in less than 1% of patients (n=1-2) and included urticaria, periorbital swelling, dyspnea, and hypoxia occurring approximately 30 minutes after administration. Other rare side effects (<5%) that may be attributable to the drug included hyperhidrosis, xerostomia, perioral hypoesthesia, and musculoskeletal pain.

##### Adverse Reactions

The data described herein reflect 2 randomized studies in patients with NHL and MM, in which 301 patients treated with Mozobil^TM^ in combination with G-CSF were compared to 292 patients who were treated with placebo and G-CSF. Side effects attributed to the drug were relatively minor and consisted of mild injection site reactions and gastrointestinal complaints (see APPENDIX D FDA-approved package insert). In particular, the most common adverse reactions (≥10%) reported in patients who received Mozobil^TM^ in conjunction with G-CSF were diarrhea, nausea, fatigue, injection site reactions, headache, arthralgia, dizziness, and vomiting.

Gastrointestinal complaints: Nausea, vomiting, and diarrhea were frequently reported in cancer patients treated with both Mozobil^TM^ and G-CSF, but also may be attributable to the chemotherapy, the apheresis, or the underlying disorder.

Injection site reactions: In the randomized studies, 34% of patients with NHL or MM had mild to moderate injection site reactions at the site of subcutaneous administration of Mozobil^TM^. This included erythema, hematoma, hemorrhage, induration, inflammation, irritation, pain, paresthesia, pruritus, rash, swelling, and urticaria.

Other adverse events: Mild to moderate systemic reactions were observed in fewer than 1% of patients approximately 30 minutes after Mozobil administration. These events included one or more of the following: urticaria, periorbital swelling, dyspnea, or hypoxia. Symptoms generally responded to treatments (e.g., antihistamines, corticosteroids, hydration, or supplemental oxygen) or resolved spontaneously (see APPENDIX D FDA-approved package insert).

##### Warnings and Precautions

**Leukocytosis**

Administration of Mozobil^TM^ in conjunction with G-CSF increases circulating leukocytes as well as HSC populations. We will monitor white blood cell counts as indicated in section 4.2.2.

**Thrombocytopenia**

Thrombocytopenia has been observed in patients receiving Mozobil^TM^. We will monitor platelet counts in all patients who receive Mozobil^TM^ as indicated in section 4.2.2.

**Splenic Enlargement and Potential for Rupture**

Higher absolute and relative spleen weights associated with extramedullary hematopoiesis were observed following prolonged (2 to 4 weeks) daily Mozobil^TM^ SQ administration in rats at doses approximately 4-fold higher than the recommended human dose based on body surface area. The effect of Mozobil^TM^ on spleen size in patients was not specifically evaluated in clinical studies. We will evaluate individuals receiving Mozobil^TM^ in combination with G-CSF who report left upper abdominal pain and/or scapular or shoulder pain for splenic integrity by physical and/or abdominal ultrasound examination.

##### Drug Interactions

Based on in vitro data, Mozobil^TM^ is not a substrate, inhibitor, or inducer of human cytochrome P450 isozymes. Therefore, Mozobil^TM^ is not likely to be implicated in drug-drug interactions involving this system in vivo.

#### Scientific and Clinical Justification

Since CXCR4 is known to be an important mediator of neutrophil retention in the bone marrow, we believe that Mozobil™ may offer a specific and well-tolerated new treatment for WHIMS and other syndromes characterized by neutropenia with myelokathexis. In the case of WHIMS, which though rare is the most common disease characterized by neutropenia and myelokathexis, it is likely that the dose needed for clinical benefit (increase in ANC to normal range) will be less than the dose already approved by the FDA for stem cell mobilization. The reason for this is that the dose used for CD34^+^ HSC harvest and HIV treatment was chosen to completely block the receptor, whereas in WHIMS our goal is simply to reduce CXCR4 function to normal levels. Thus, our primary goal in this protocol is to provide proof of principle that Mozobil™ can be used to increase the ANC in patients with neutropenia and myelokathexis to levels that are not associated with an increased risk of infection. Exploratory goals of the study are to determine whether Mozobil™ administration increases other leukocyte subsets in the peripheral blood, and to determine whether HPV lesions are ameliorated.

#### Qualifications of Investigators

This protocol will be conducted by a multidisciplinary team with expertise in the area of immunology, infectious diseases, pharmacokinetics, and dermatology. The care team is also knowledgeable in the conduct of GCP principles of clinical research and the regulatory requirements for the protection of human subjects. All investigators collaborating in this study have met the training requirements of the Office of Human Subjects Research. Copies of curriculum vitae to demonstrate the experience and qualification of all of the investigators (PI and Associate Investigators [AIs]) will be kept updated and on file.

#### Conflict of Interest

All investigators of this study will be asked to complete the NIH financial disclosure form. Currently, no reportable conflicts of interest have been identified for any of the investigators who will be conducting this protocol. If conflicts of interest should arise, the IRB will be notified.

#### Conduct of the Protocol

This protocol will be conducted in accordance with all applicable laws regulations, and policies of the NIH IRB as well as the policies of NIAID and NIH. The PI will assure that no deviation from or changes to the protocol will take place without prior documented approval from the IRB, except where necessary to eliminate an immediate hazard(s) to the protocol participants. The PI will promptly report to the IRB any changes in research activity and all unanticipated problems involving risk to human subjects or others.

#### Observed Risks and Incidents in an NIH Study with Plerixafor and G-CSF

In another NIH study (14-I-0185) where patients receive two separate treatments of 14 months plerixafor and 14 months of G-CSF in random sequence and patients and study staff are not informed of which drug is being administered,19 WHIM patients have been enrolled and have received both study drugs. A subject experienced a new severe rash (psoriasis-like) beginning a week after commencing the 1^st^ study drug, which resolved after switching to the alternate study drug and topical steroid treatment. Another subject who has never received G-CSF prior to the study experienced pain and swelling in his hands and knees on G-CSF and on each of the study drugs and was discontinued from both study agents. The subject was diagnosed with arthralgia and reactive arthritis and his symptoms resolved with oral steroid treatment.

### STUDY DESIGN

#### Overview

The objectives of this study are to determine whether or not Mozobil^TM^ is safe in this patient population, whether the drug is effective at increasing the ANC, and to understand the pharmacokinetics of the drug in this patient population. This study consists of 4 phases which may be performed sequentially or separately: phase A, phase B, phase C and phase D. Subjects who volunteer for this study will have an initial screening evaluation and interruption of their chronic therapy (G-CSF/GM-CSF washout phase; see Section 4.2.1 #14) before admission to the NIH Clinical Center for a drug escalation study to determine a dose that increases patient ANC to 4000 cells/microliter of blood (Phase A, Study Drug Escalation phase schedule in Section 4.2.2). For safety reasons and to determine the optimum dose in this patient population, the drug will be slowly increased (dose escalation) until normalization of neutrophil counts (ANC greater than 4000 cells/microliter of blood) or until the maximum FDA daily dose is achieved. When this dose is determined or the maximal FDA-approved dose is reached, we will administer that same dose 48 hours later to study drug pharmacokinetics and circulating neutrophil kinetics (Phase B, Response to Maximum Dose phase, Section 4.2.3). Phase B is designed to deliver a single dose of the final dose defined in phase A or the FDA-approved dose in order to be able to define drug pharmacokinetics in this patient population and to relate it to the kinetics of the neutrophil and HSC response. Pharmacokinetics of the drug will be determined by frequent sampling of plasma after the dose followed by laboratory determination of drug levels coupled with frequent monitoring of the ANC. Phase C allows patients to receive the drug via syringes for up to 84 months, while Phase D is a trial to deliver the drug via a disposable infusion pump for up to 12 months.

##### Phase A, B, or A & B: Dose Escalation (optional)

If required to determine the MED, a dose escalation study may be conducted at the discretion of the PI. In the dose escalation Phase A, B, or Phases A and B, saline or drug will be administered SQ once daily (QD) by qualified personnel using an insulin type syringe and needle using proper sterile technique. The dose will be determined in consultation with the NIH Clinical Center Pharmacy and the administration of the drug will be monitored and recorded electronically in Clinical Research Information System (CRIS) by trained personnel. Each participant will have a baseline electrocardiogram (EKG) and echocardiogram (ECHO) prior to initiation of the study drug. In addition, baseline safety laboratory tests will be conducted on blood samples. During the dose escalation phase, a CBC test and CHEM7 test panel will be determined on patients prior to each dose and safety tests will be conducted each day the participant is on drug. In addition, an SQ injection of normal saline equivalent in volume to the volume of the final drug dose will be given followed by blood draw 6 hours later for CBC.

##### Phase C: Chronic Dosing with Syringes (optional)

Patients may opt to do up to 84-month Phase C (chronic dosing with syringes). During this phase, G-CSF/GM-CSF will be held and Mozobil^TM^ will be given twice a day. The maximum allowable daily plerixafor dose in this phase is 0.08 mg/kg. The initial dose will be one half the MED determined in phase A given twice per day, or if a recent phase A was not conducted, then the dose will be approximately 0.03 mg/kg per day or the closest syringe dose available from pharmacy. The dose will be adjusted to maintain adequate leukocyte response. After the first 3 months, with the PI’s approval, patients may opt to switch to daily injections. After the initial switch, in consultation with the PI and planning for study drug supplies, patients are permitted to switch between daily or twice daily injections. Each participant will have a baseline EKG and ECHO prior to initiation of the study drug. There will be optional bone marrow biopsies, wart biopsies, and wart photographs for research purposes. The Schedule of Events for Phases C & D is described in APPENDIX F.

##### Phase D: Trial Using an Infusion Pump – in place of syringes (optional)

Patients may participate in Phase D, a trial with plerixafor infused with disposable infusion pumps instead of syringes. G-CSF/GM-CSF will be held and Mozobil^TM^ will be infused through a pump, either as a bolus, continuous infusion, or a combination of both, for a maximum of 24 months and with a total infused dose not to exceed the study’s daily maximum of 0.08 mg/kg. The initial dose will be approximately 0.03 mg/kg per day and adjusted to maintain adequate leukocyte response. Each participant will have a baseline EKG and an ECHO within the last 5 years prior to initiation of the study drug. There will be optional bone marrow biopsies, wart biopsies, and wart photographs for research purposes. The Schedule of Events for Phases C & D is described in APPENDIX F.

###### Infusion Pump: Training, Monitoring, and End of Trial

Use of the pump requires additional training to operate, dose, and monitor response as described in APPENDIX H:

1. Training (Day -1): patient demonstrates ability to operate pump – loaded with saline.
2. Start of study drug (Day 0): patient operates pump containing the study drug plerixafor.
3. Measure response to bolus infusion (Day 0, first 12 hours).
4. Measure response to start of continuous infusion (Day 0-1).
5. Adjust dose to achieve desired leucocyte response (Day 1-2).
6. Ensure patient is trained and has pump supplies prior to discharge (Day 2-3).
7. Monitor leukocyte response at home (Weeks 2 & 4).
8. NIH interim visit (Month 3 ± 1 month).
9. Monitor leukocyte response at home (Months 6 & 9 ± 1 month).
10. NIH end of trial visit (Month 12 ± 1 month).

#### Study Endpoints

##### Primary Endpoints

The primary endpoints for this study are the following:

1. Safety in this patient population defined as no occurrences of Grade III/IV toxicities that require temporary hold of the study drug.
2. Efficacy in this patient population, defined as: i. in Phase A, determination of whether treatment with Mozobil^TM^ is effective at raising the ANC at least 2-fold over the baseline average pretreatment value; and ii. In Phase C and D, determination of whether treatment with Mozobil^TM^ is effective (measured just before drug dosing for Phase C) at raising the ANC to BOTH an average >250 cells/μL AND at least a 2-fold increase over baseline average pretreatment value.

##### Exploratory Endpoints

The exploratory endpoints for this study are the following:

1. Improvement in other leukocyte subsets—Lymphocyte phenotyping will be performed on Day −1 or Day 0 before initial drug administration as a baseline and once after the last dose of drug administered. Success will be defined as a significant increase in cell number or percent after drug administration.
2. Reduction in the size of HPV lesions—With subject consent, warts on the skin and oral, nasal, and genital mucosa will be photographed prior to initiating drug and compared to those taken on the last dose of drug. Wart biopsies may be taken before and after drug dosing to look for viral persistence or clearance from the lesions.
3. Immune response to treatment**—**We expect WHIM patients’ immune system to exhibit changes when CXCR4 hyperfunction is treated; therefore, we will study a variety of cytokines, chemokines, their receptors, specific antibody responses to vaccination, and other leukocyte functional studies as appropriate.

#### Sample Size Justification

In view of the absence of reported studies for the use of Mozobil^TM^ in the treatment of myelokathexis, the sample size for this study will be 20 patients. However, we will obtain safety and efficacy data on the first patient; results of which will be evaluated to determine whether subsequent subjects will be enrolled into the study. Upon completion of the first patient to receive Mozobil^TM^, safety data will be submitted to IRB for review.

### SUBJECT ENROLLMENT

#### Patient Recruitment

Participants in this protocol will be recruited from patients with documented severe infection who have WHIMS, particularly those who are intolerant or who have side effects from the FDA‑approved standard of care medication for all patients with SCN, G-CSF (Neupogen, Zarxio or other biosimilar agents) or who have major clinical issues precluding participation in a longer‑term blinded trial of plerixafor and G-CSF such as protocol 14-I-0185. Recruitment will be accomplished by public announcement to primary care providers, dermatologists, and allergists. The Clinical Center Patient Recruitment and Public Liaison Office serves to provide protocol advertisement and contact information for both self-referring patients and physician referrals from outside NIH.

Because the length of Phase C has been extended from 6 months in the 2010 version to 84 months in the 2021 version, individuals who participated in the original trial will be eligible to re-enroll in this extended Phase C study or in the Phase D study. Phase D is a 24-month trial with a maximum of 10 participants, to deliver the drug via an infusion pump in place of syringes; eligibility for this phase is similar to Phase C.

#### Inclusion/Eligibility Criteria

All of the following inclusion criteria must be met for a subject to be enrolled in this study:

1. Clinical diagnosis of WHIMS and documented severe infection.
2. Must be ≥18 and ≤75 years of age.
3. Willingness to interrupt medications to raise the WBC such as G-CSF or GM-CSF for at least 2 days before and while on the study drug.
4. Must not be pregnant or breastfeeding.
5. Must have a personal physician.
6. Must be willing to provide blood, plasma, serum, and DNA samples for storage.
7. Subjects must agree not to become pregnant or to impregnate a partner. If of childbearing potential, must agree to consistently use two types of contraception throughout study participation. Acceptable forms of contraception include the following:
   a. Condoms, external or internal, with or without a spermicide.
   b. Diaphragm or cervical cap with spermicide.
   c. Intrauterine device.
   d. Contraceptive pills or patch, Norplant, Depo-Provera, or other FDA-approved contraceptive method.
   e. Partner has previously undergone a vasectomy for which there is documentation of aspermatogenic sterility.

#### Exclusion Criteria

If any of the following exclusion criteria are met, a subject will not be enrolled in this study:

1. Absence of a diagnosis of WHIMS.
2. Patient is less than 18 years old.
3. Absence of a documented history of severe infection.
4. Neutropenia due to maturation defects in the myeloid lineage or that the PI feels is unlikely to benefit from this medication.
5. Pregnant or breastfeeding.
6. History of serious cardiac arrhythmia or cardiac defects that make such more likely.
7. Renal failure (calculated creatinine clearance [CrCl] <15 mL/min or requiring dialysis).
8. Signs or symptoms of active microbial infection at the time of study entry.
9. Any condition that, in the investigator’s opinion, places the patient at undue risk by participating in the study.
10. Unwillingness to undergo testing or procedures associated with this protocol.

#### Study Enrollment Procedures

##### Informed Consent

Informed consent is a process that is initiated before an individual agrees to participate in the study and continues throughout the individual’s study participation. Extensive discussion of risks and possible benefits of this therapy will be provided to the participants and their families. Consent forms will be approved by the IRB, and the participant will be asked to read and review the document.

The informed consent document will be provided as a physical or electronic document to the participant or consent designee as applicable for review prior to consenting. A designated study investigator will carefully explain the procedures and tests involved in this study, and the associated risks, discomfort, and benefits. In order to minimize potential coercion, as much time as is needed to review the document will be given, including an opportunity to discuss it with friends, family members, and/or other advisors, and to ask questions of any designated study investigator. A signed informed consent document will be obtained prior to any research activities taking place.

The initial consent process as well as re-consent, when required, may take place in person or remotely (e.g., via telephone or other NIH-approved remote platforms used in compliance with policy, including Human Research Protection Program [HRPP] Policy 303) per the discretion of the designated study investigator and with the agreement of the participant. Whether in person or remote, the privacy of the subject will be maintained. Consenting investigators (and participant, when in person) will be located in a private area (e.g., clinic consult room). When consent is conducted remotely, the participant will be informed of the private nature of the discussion and will be encouraged to relocate to a more private setting if needed. If the consent process is occurring remotely, participants and investigators will view individual copies of the approved consent document on screens at their respective locations; the same screen may be used when both the investigator and the participant are co-located.

Note: When required, the witness signature will be obtained similarly as described for the investigator and participant below.

Consent will be documented with required signatures on the physical document (which includes the printout of an electronic document sent to the participant) or on the electronic document. The process for documenting signatures on an electronic document is described below.

When a hand signature on an electronic document is used for the documentation of consent, this study will use the iMedConsent platform (which is 21 CFR Part 11 compliant). Both the investigator and the participant will sign the electronic document using a finger, stylus, or mouse. Electronic signatures (ie, the “signature” and a timestamp are digitally generated) will not be used.

The acquisition of informed consent will be documented in the participant’s medical records, as required by 21 CFR 312.62. The informed consent form will be signed and personally dated by the participant and the person who conducted the informed consent discussion. The original signed informed consent form will be retained in the medical chart and a copy will be provided to the participant.

At any time during participation in this protocol that new information becomes available relating to risks and/or AEs, this information will be provided orally and/or in writing to all enrolled or prospective patient participants. Documentation will be provided to the IRB and if necessary, the informed consent will be amended to reflect relevant information.

##### Justification for Exclusion of Women, Minorities, and Children (Special Populations)

Children: Although myelokathexis is a rare congenital immunodeficiency that affects all age groups, in the absence of reported studies for the use of Mozobil^TM^ in the treatment of WHIMS, this study will only include adults.

Pregnancy: Pregnant people are excluded from this study because the effects of Mozobil™ (plerixafor injection, Genzyme/Sanofi) on the developing human fetus are unknown with the potential for teratogenic or abortifacient effects.

##### Request for Waiver of Informed Consent for Secondary Research from NIH IRB Protocol 14-I-0185

The informed consent form for NIH protocol 14-I-0185 (see Section 5) stated that participants’ coded samples with the code key, as well as data including health history, age, and race, may be shared with other investigators.

We request a waiver of informed consent for this secondary research, consistent with the pre‑2018 45 CFR 46.116 (d):

1. The research involves no more than minimal risk to participants:
   a. The only risk of this secondary research is a breach of confidentiality. Confidentiality protections are described in section 8.4.
2. The research could not practicably be carried out without the requested waiver:
   a. The original protocol is now closed and we no longer have contact with the previous participants.
3. The research could not practicably be carried out without using the biospecimens and data in an identifiable format:
   a. The identifiable information is required to link the corresponding data to the biospecimens.
4. The waiver will not adversely affect the rights and welfare of the participants:
   a. We do not intend to have any contact with the participants whose biospecimens or data we will use in this study.

### STUDY IMPLEMENTATION

#### Criteria for Withdrawal

##### Voluntary Withdrawal

All subjects have the right to voluntarily discontinue participation in the study at any time. Study subject will be removed from the protocol and at the PI’s discretion may be replaced for any of the following reasons:

- If a participant refuses study drug before reaching the primary endpoint during any point in the Active phase (see section 4.2.2).
- If a participant experiences Grade III/IV toxicity that is not related to study drug.

##### Involuntary Withdrawal

A study subject may be withdrawn without their consent for any of the following reasons:

- Patient develops any of the excluded medical conditions (see Section 3.3).
- If this clinical trial is officially terminated. This applies to the situation where the investigator for any reason, terminates the entire study.
- If the subject has a serious adverse event (SAE), classified as possibly, probably, or definitely related to Mozobil^TM^, the administration of the drug will be discontinued, and the patient will be terminated from the study. Follow-up visits (frequency determined by treatment regimen for the SAE) will be required.
- Therapy will be stopped if a patient requires treatment with a prohibited medication (see Section 3.3).
- A subject becomes pregnant.
- A subject is noncompliant with requirements and procedures of the study.

#### Study Parameters

Subjects will be screened for suitability for this protocol using an existing protocol such as the NIAID Screening Protocol (05-I-0213) for Host Defense Defects for the determination of suitability for this trial. The purpose of the pre-drug screening in this protocol is to further determine volunteer eligibility for study participation and ensure patient safety and act as a baseline for AEs. Subjects who are diagnosed with a medical condition during the screening process (e.g., test positive for hepatitis B, hepatitis C, or HIV) will be notified and referred for medical care with their personal care provider.

##### Pretreatment Evaluations Within 30 Days

The study will comprise a baseline run-in period. Candidate subjects will initially be interviewed to confirm their diagnosis of WHIMS (clinical history and supporting data). This can be done using outside reports from a referring physician or by evaluation at the Clinical Center. Once a diagnosis of WHIMS is confirmed, subjects will be seen for a screening visit in the NIH Outpatient Clinic for the following:

1. The informed consent forms will be reviewed and signed.
2. Complete history and physical examination.
3. Blood tests will include hematology (including CBC and platelet count), coagulation panel (including prothrombin and partial thromboplastin times [PT/PTT]), erythrocyte sedimentation rate (ESR), chemistry 20 panel (including acute care, mineral, and liver function panels), vitamin B12, folate, and C-reactive protein.
4. Human lymphocyte antigen (HLA)-type (A, B, Cw, DR, and DQ).
5. Immunological studies: lymphocyte subsets (lymphocyte phenotyping B cell panels 1 and 2), quantitative immunoglobulins, complement studies, lymphocyte proliferation studies, functional studies such as dihydrorhodamine testing, and antibody levels for diphtheria and tetanus.
6. *HIV antibody test, hepatitis A antibody, B surface antigen, hepatitis C antibody.
7. Serum pregnancy test for participants of childbearing potential.
8. Urinalysis determinations of blood urea nitrogen and creatinine.
9. EKG and ECHO to rule out cardiac problems.
10. Baseline spleen measurement via ultrasonography.
11. Photograph of wart areas if patient signs a separate medical photography consent.
12. Optional baseline wart biopsy for research purposes if participant consents.
13. Research bloods samples will include the following: 5 mL of EDTA anticoagulated blood (1 purple top), 5 mL of serum (1 tiger top), and 30-60 mL of heparinized blood (3-6 green tops) will be collected for DNA/RNA isolation, EBV-transformed cell line creation, serum/plasma storage, and flow cytometry.
14. Drug washout for patients GM-CSF/G-CSF as follows:

| Day | Event |
| --- | --- |
| Day -2 | - Stop GM-CSF and/or G-CSF injections - Patient 2-day symptom diary |

Patients who do not fulfill the criteria on screening will be discharged back to the care of referring physicians.

* These tests may not need to be repeated if done within 2 months on another NIH-approved protocol prior to enrollment onto this protocol.

##### Active Phase A (Dose Escalation Phase Inpatient)

All patients will be admitted 2 days prior to the initial Mozobil^TM^ injection. We anticipate an inpatient stay of less than 14 days. All patients will have the following performed before the first injection:

- A directed history and physical examination.
- Symptom and medication log review.
- Urinalysis, urine/serum pregnancy test, PT/PTT, chemistry 20 panel, ESR, lymphocyte phenotyping B cell panels 1 and 2, quantitative serum immunoglobulin levels.
- Daily safety laboratory tests: CBC, acute care panel.
- Saline injection test at the minimal volume (equivalent volume to the 0.02 mg/kg dose SQ) on Day -1.

Subjects will receive Mozobil^TM^ as follows:

**Study Drug Escalation Phase A Schedule**

| **DAY** | **DOSING**** | **EVENT** |
| --- | --- | --- |
| Day-2 | None | - CBC at 9am, 12pm, 3pm, 6pm. QD acute care, ESR, urine test |
| Day -1 | Normal saline (at volume equivalent to the 0.02 mg/kg dose) injection test | - Pre-injection of saline test. CBC at 9am, 12pm, 3pm, 6pm, QD acute care, ESR |
| Day 0 | 0.02 mg/kg | - Pre-injection: obtain CBC, QD acute care, ESR - Post-injection: Vital sign 30 mins (±10 mins), 60 mins (±10 mins), CBC at 3, 6, and 9 hours after injection - Complete medication/symptom log |
| Day 1 | 0.04 mg/kg | - Pre-injection: obtain CBC, QD acute care, ESR - Post-injection: Vital sign 30 mins (±10 mins), 60 mins (±10 mins), CBC at 3, 6, and 9 hours after injection - Complete medication/symptom log |
| Day 2 | 0.08 mg/kg | - Pre-injection: obtain CBC, QD acute care, ESR - Post-injection: Vital sign 30 mins (±10 mins), 60 mins (±10 mins), CBC at 3, 6, and 9 hours after injection - Complete medication/symptom log |
| Day 3 | 0.16 mg/kg | - Pre-injection: obtain CBC, QD acute care, ESR - Post-injection: Vital sign 30 mins (±10 mins), 60 mins (±10 mins), CBC at 3, 6, and 9 hours after injection - Complete medication/symptom log |
| Day 4 | 0.24 mg/kg | - Pre-injection: obtain CBC, QD acute care, ESR - Post-injection: Vital sign 30 mins (±10 mins), 60 mins (±10 mins), CBC at 3, 6, and 9 hours after injection - Complete medication/symptom log |
| Day 5 | None | Obtain CBC at 9am, 12pm, 3pm, 6pm; QD acute care, ESR |

**Dose increase occurs if Absolute Neutrophil Count (ANC) is less than 4000 cells/microliter of blood and previous dose has been tolerated without Grade III/IV toxicities. Dose is calculated based on actual body weight and it is reduced by 1/3 if CrCl <50 mL/min and is given sq once daily but should not exceed 40 mg/day (27 mg/day if renal impairment exists).

##### Active Phase B (Response to Maximum Dose - Inpatient)

**Schedule**

| **DAY** | **DOSING**** | **EVENT** |
| --- | --- | --- |
| Day 0 | 0.24 mg/kg | - Pre-injection: obtain CBC, 10 mL purple top, acute care, ESR - Post-injection: Vital signs 30 mins (±10 mins), 60 mins (±10 mins), CBC and 10 mL purple top for HSC measurement at 3, 6, 9, 12, 24 hours after dose. - Complete medication/symptom log - Obtain EKG, send immunophenotyping B cell panels 1 and 2, quantitative immunoglobulin levels if not participating in phase C. |
| Day 1 | normal saline equivalent in volume to the volume of the final drug dose will be given by subcutaneous injection as a vehicle control | - Post-injection of saline: blood draw 6 hours later for CBC - Get another heart tracing (EKG). |

**Dose increase occurs if Absolute Neutrophil Count (ANC) is less than 4000 cells/microliter of blood and previous dose has been tolerated without Grade III/IV toxicities. Dose is calculated based on actual body weight and it is reduced by 1/3 if CrCl <50 mL/min and is given sq once daily but should not exceed 40 mg/day (27 mg/day if renal impairment exists).

##### Chronic Phase with Syringes (Phase C)

Patients have the option to participate in a chronic phase for a total of up to 84 months. Patients may begin either directly after phase A/B is completed or be started separately later. During this chronic phase, regardless of the method of entry, patients return to NIH at the end of month 3, 6 (± 1 month) then every 3 or 6 months (± 1 month) thereafter.

**Direct Entry to Phase C**

The chronic dose (CD) for Phase C is one-half of the MED. MED is determined in Phase A as the dose at which the ANC BOTH doubles relative to the pre-drug average baseline value AND exceeds 500 cells/μL.

| **DAY** | **DOSING** | **EVENT** |
| --- | --- | --- |
| Day 9 or 5^th^ day after last phase A dose | CD Q12h | CBC before and at 3 hours after AM injection. Optional CBC at 3, 6 and 9 hours after AM injection |

**Indirect Entry to Phase C**

The CD for Phase C is one-half of the MED. MED is determined in Phase A as the dose at which the ANC BOTH doubles relative to the pre-drug average baseline value AND exceeds 500 cells/μL. If Phase A was not conducted and after two weeks of chronic treatment, the nadir ANC has not risen above 250 neutrophils/microliter of blood, then the dose will be increased as required and if it remains below 250 neutrophils/microliter of blood after 3 months, and this is repeated within one week, then the phase will be halted and the patient will be considered to have failed therapy.

| **DAY** | **DOSING** | **EVENT** |
| --- | --- | --- |
| **Day-2 to start of Phase C** | None | Stop GM-CSF and/or G-CSF injections at least 2 days before Phase C initiation. |
| **Day 0** | MED Q12h | CBC before and at 3 hours after AM injection. Optional CBC at 3, 6 and 9 hours after AM injection. |

##### Pilot Trial with Infusion Pump (Phase D)

Patients have the option to participate in a trial to receive the study drug via an infusion pump (instead of syringes) for a total of up to 24 months, as described in APPENDIX H. Patients not on immunoglobulin supplementation have the option of receiving pneumococcal vaccination during the 24-month period in order to measure their IgG response with continuous plerixafor infusion.

##### Phase C and D Safety Monitoring

Per the schedule of events in APPENDIX F, the following safety monitoring will be done:

- CBC and acute care panel for Weeks 2 and 4 (± 1 week) and at months 2 & 3 (± 2 weeks) and (just before dosing for Phase C), then every 3 months to the end of the study.
- Immunoglobulin levels (IgG, IgA, IgM, and IgE) will be measured at NIH visits. Immunophenotyping will be conducted as required.
- Optional bone marrow biopsies and wart biopsies may be performed annually or prior to drug completion. Photographs of warts and skin lesions will be taken as applicable at NIH visits.

##### Study Completion Visit

A study completion visit will be conducted 6 months (±2 months) of the last dosing of Mozobil^TM^. At this outpatient visit, the following will be assessed:

- A directed history and physical examination.
- Symptom and medication log review.
- Urinalysis, urine/serum pregnancy test, PT/PTT, chemistry 20 panel, ESR, CBC, lymphocyte phenotyping B cell panels 1 and 2, quantitative serum immunoglobulin levels.
- Optional bone marrow biopsy, wart biopsy and wart photographs.

At the end of the study completion visit, subjects will be returned to the care of their local physicians.

#### Collaboration with Other Investigators

Coded biological samples may be sent to outside investigators who will not be provided with the key to the code, and thus will not be engaged in human subjects research. Results of these assays may be returned to the NIH study team to be relinked to the participants if deemed necessary for further analysis.

This protocol will also serve as a secondary research use protocol for stored biological samples collected in NIH IRB protocol 14-I-0185. The purpose of that study, closed as of November 2023, was to compare severity of infections during treatment with either plerixafor or G-CSF in people with WHIM syndrome. Blood counts and immunologic parameters were secondary endpoints.

This may improve our understanding of how these interventions affected the participants. Conducting secondary research on this protocol simplifies the logistics and operational concerns of data and sample sharing. All participants of NIH protocol 14-I-0185 provided informed consent for storage and sharing of data and samples for future research, and thus do not need to provide additional informed consent (section 3.4.3).

#### Failure to Respond to Mozobil^TM^

**In Phase A, failure** will be defined as the inability to achieve a 2-fold increase of ANC over baseline average pre-drug ANC by the maximum dose of plerixafor given. In Phase B, a single additional dose to establish the pharmacokinetics of the drug in this patient group, will be performed at the dose needed to obtain an ANC of 4,000 cells/microliter or at the 0.24‑mg/kg dose.

**In Phases C and D, failure** will be defined in two ways:

- If by the 3-month visit, ANC at nadir (predicted trough drug level in Phase C or Phase D) remains below 250 neutrophils/microliter of blood at a dose of at least 0.08 mg/kg per day and this value is confirmed by repeat nadir CBC within one week. Patients failing by this criterion will not be eligible for further drug administration in this protocol.
- The average value of all available nadir ANC’s (predicted trough drug level in Phase C or Phase D) on drug remains <250 neutrophils microliter of blood OR <2-fold greater than the baseline average pre-drug value.

#### Dose/Schedule Modifications for a Participant

Each participant will be given study drug as indicated in the dose escalation schedule unless the study endpoint is reached or there is a Grade III/IV toxicity that is possibly, probably, or definitely related to the study agent. The participant may refuse study drug, but such refusal will result in removal from the protocol.

In Phase C and D, dosing changes will depend on neutrophil and lymphocyte levels. The dose may be increased up to the maximum allowable dose, and if it remains below 250 neutrophils/microliter of blood at the 3-month visit, and this is repeated within one week, then the phase will be halted, and the patient will be considered to have failed therapy. If on the other hand at any time the patient’s ANC rises to >4000 in the absence of signs of clinical infection, the dose will be reduced.

In phase D, when using the infusion pump to deliver plerixafor, the subject will be trained with saline infusion prior to start of the study drug, receive a one-time bolus of plerixafor on the first day, and commence continuous infusion later that day or the next day. The ANC and absolute lymphocyte count (ALC) will be monitored, and the dose adjusted to ensure a minimum dose response is achieved prior to the patient leaving the NIH. The pump controller will be programmed with doses of plerixafor in increments of approximately 0.01 mg/kg/day and infused at a rate to account for a mean half-life of 5.2 hours, clearance of 5.2 L/h and volume of distribution of 30.2 L.[12] Plerixafor infusions at the NIH may include a loading dose to achieve steady state drug concentration sooner thereby allowing the study team to monitor dose response sooner and adjust the dose while the patient remains at the NIH. At home, the patient’s blood counts will be monitored at weeks 2 and 4, and if necessary, the dose can be adjusted in coordination with the study team, by the patient selecting alternate preprogrammed doses from their controller. A record of the doses and infusion amounts is recorded in the pump history and will be collected and reviewed by the study team (APPENDIX G, APPENDIX H).

#### Study Drug Acquisition

Mozobil™ is FDA-approved and manufactured by Pantheon UK Ltd, Swindon, UK for Genzyme/Sanofi, Cambridge, MA, USA. Drug will be provided by the manufacturer and distributed through the NIH Clinical Center Pharmacy Department. Remaining study medication will be returned to the NIH Clinical Center Pharmacy Department or destroyed.

#### Prohibited Medications and Procedures

Participants are not to receive medications that raise the WBC such as G-CSF or GM-CSF during this study. All pre-protocol medications will be re-started at the end of active phase A or B unless patient elects to participate in optional Phase C and/or D. Patients will continue to receive any prophylactic antibiotics that they have been receiving prior to participation on this study. If they have a documented infection during phase C or D period despite this, the PI should immediately be contacted and if the ANC is <250, G-CSF will be administered, and the phase C or D will be halted, and appropriate antibiotics given. If the ANC is >250, appropriate antibiotics will be administered and the Mozobil will be continued.

#### Subject Monitoring and Supportive Care

Patients will be closely monitored for any medication side effects or toxicities. The common toxicity criteria (CTC version 3.0 available at <http://ctep.cancer.gov/protocolDevelopment/electronic_applications/ctc.htm>, specifically the applicable subsections that describe the following toxicities: Allergy/Immunology, Constitutional Symptoms, Death, Pain, Blood and Bone Marrow, Neurological, Musculoskeletal, Gastrointestinal, Dermatology/Skin and Infection) will be used to classify and grade adverse events (see APPENDIX A). The data will be collected on case report forms for serious and non‑serious AEs. Each patient will be provided contact information that allows direct and immediate access to the Dr. McDermott, the PI for trial information, and recording of AEs.

**Rescue Medications**

In the event of a severe vasovagal or anaphylactic reaction, standard medical therapies will be employed if necessary. Injection site reactions usually resolve rapidly without treatment, but topical corticosteroids or oral antihistamines may be helpful. Gastrointestinal side effects will be treated symptomatically.

#### Temporary Suspension Rules for the Protocol

The PI will closely monitor and analyze study data as they become available and will make determinations regarding the presence and grading of AEs. Evaluation of AEs will be analyzed for Mozobil™ with regard to the known complications associated with administration. The following events will result in the temporary hold of this protocol:

- If Grade III/IV toxicities are found to be definitely related to study drug.
- If infection develops per sections 6.1 and 6.2.
- Evidence of severe splenic enlargement on imaging study that is possibly, probably, or definitely related to study drug.

Upon the discovery of any of the above, the study will be placed on temporary hold (no new enrollments and no further administration of Mozobil™ by the investigators) and a report will be submitted to the NIH IRB. The study may resume after consultation and approval by NIH IRB.

The NIH IRB as part of their duties to ensure that research subjects are protected; may discontinue the study at any time. Subsequent review of serious, unexpected, and related AEs by the NIH IRB may also result in suspension of further trial interventions/administration of study agent at a site.

#### Stopping Rules for an Individual Participant/Cohort

A study participant will be discontinued from further study agent(s)/intervention(s) administration for:

- Any clinical AE, laboratory abnormality, intercurrent illness, other medical condition, or situation occurs such that continued participation in the study would not be in the best interest of the participant.
- Development of any exclusion criteria may be cause for discontinuation.
- Phase C and D – failure to achieve nadir ANC >250 after 3 months of treatment.

### ANALYSIS OF STUDY

This is a pilot study of the effectiveness of Mozobil^TM^ in patients with WHIMS. Therefore, each patient in this study will serve as their own control using comparisons of laboratory values and symptoms and signs before and after drug administration. Analysis of the first primary endpoint—safety—will be based on a yes/no determination of safety as defined by no occurrence of Grade III or Grade IV toxicities during the study. Safety and side effects will be assessed with the safety tests and case report form. Analysis of the second primary endpoint—efficacy—will be a yes/no determination based on an increase of the ANC by 2-fold over pre-drug levels (average of at least 2 CBC tests performed at the same time of day to control for diurnal variation) in phase A. The drug will be considered ineffective if there is not at least a 2-fold increase of the ANC at the highest dose in phase A. A paired t-test will also be used to compare before and after drug administration ANC values in the same patient and a p value of 0.05 will be considered significant. For safety reasons in Phase A, the drug is being initiated at a low dose and then increased until the maximum approved daily dose is reached, or ANC is normalized to a value of >4000 cells/microliter. The optimum dose of Mozobil^TM^ will be that which increases the ANC by at least 2-fold and in the judgment of the PI causes a minimal number of side effects and no Grade III or Grade IV toxicities. In Phase B, we will sample blood frequently in order to create a pharmacokinetic curve of drug levels achieved and a comparison with circulating white count using appropriate dynamic modeling. It is expected that the pharmacokinetic curve will be similar to that already obtained in healthy normal subjects; however, we wish to study how the white blood cell and HSC count responds and with what time delay to a particular drug level in order to determine optimum long-term dosing. In phase C and D, safety will be defined exactly as in phase A/B; however, analysis of the second primary endpoint, efficacy will be a yes/no determination based on an average nadir ANC >250/µL over the entire treatment period without the addition of G-CSF or other hematopoietic growth factors and with the caveat that all individuals who fail to reach a nadir ANC >250/µL by 3 months of chronic treatment on two separate measurements will be considered efficacy failures.

The goal of phase D is to evaluate the performance of continuous delivery of Mozobil^TM^ via pump over 24 months and to compare a day of continuous infusion versus a day of bolus infusion in terms of blood counts. Previous data has shown that a bolus infusion of Mozobil^TM^ sharply increases blood counts after a short lag followed by a rapid decline. Based on three patients, we calculated the maximum less the minimum of ANC counts at 0, 3, 6, 9, and 24 hours post-infusion also known as the range. For the three patients, the mean log range was 2.75 with a standard deviation of 0.20. Assuming that the log-range is cut in half on continuous infusion, and that the standard deviation of the difference in log range is 0.20, we have excellent power (>99%) to detect a difference in log range between the day of bolus infusion compared to the day of continuous infusion. This calculation is based on a one sample two-sided t-test with alpha=0.05 for the difference in log range with 4 subjects. We conservatively use 4 subjects even though our goal is to enroll 5. Power for other blood counts (ALC, absolute monocyte count, WBC) is even greater than for ANC. To assess long term performance of continuous infusion of Mozobil™ we will describe blood counts over time (week 2, week 4, months 2, 3, 6, 9, and 12) by spaghetti plots and linear regression. The expectation is that these counts will be relatively large (for this population) and relatively stable. We will also measure IgG over time (month 0, month 3, and month 12), and describe IgG change using spaghetti plots and linear regression. The expectation is that IgG may increase over time. Any IgG values that are contemporaneous with IVIG administration will be omitted for these analyses. We will also descriptively assess how warts change over time by taking pictures of relevant regions at month 0, month 3, and month 12 and a 50% decline in wart area will be considered significant.

Exploratory endpoints of the study will include a lymphocyte phenotyping assay and quantitative immunoglobulin determinations to be done once pre-drug, once every 3 months during chronic drug treatment, and once post-drug. We will compare particular subtypes of other leukocytes (monocytes, B cells, CD4+ and CD8+ T cells, etc.) and particular types of immunoglobulins (IgG, IgA, IgM, etc.) and measurement of B cell class switching to see how these respond as well and again a paired t-test with will be used for comparison and a significance level of p=0.05 will be used in the analysis.

If a patient withdraws from this study prior to completion of Phase A due to any reason other than a Grade III or IV toxicity, their data will not be included in the Phase A analysis. If a participant does have to withdraw from Phase A because of a Grade III or Grade IV toxicity, their data from the first doses may be included if the blood sampling was complete for that dose per the protocol. Similarly, if a patient fails to complete the blood sampling or get the dose administered in Phase B, they will be excluded from the Phase B analysis. If a patient agrees to participate in phase C or D but fails to complete the maximum period of treatment because of a voluntary withdrawal, their data up until the time of withdrawal will still be used in the analysis. Therefore, the analysis of this study will not be on an “Intention to Treat” basis but rather an actual treatment basis.

### POTENTIAL HAZARDS AND DISCOMFORTS

#### Potential Risks of Mozobil^TM^

**Fetal harm in pregnancy:** Human fetal risks associated with Mozobil™ are not known, but pre‑clinical animal data have indicated that there may be some risk. Therefore, subjects must agree not to become pregnant or to impregnate a partner. Participants of childbearing potential must have a negative pregnancy test before beginning the study agent. Because of the risk involved, subjects and their partners must use 2 methods of birth control. They must continue to use both methods until 1 month after stopping the study drug. Two of the birth control methods listed below may be chosen:

- Hormonal contraception
- External or internal condoms with or without a spermicidal
- Diaphragm or cervical cap with a spermicidal
- Intrauterine device (IUD)

Patients who become pregnant on study will be followed to term for safety monitoring.

**Breastfeeding:** Because there is an unknown but potential risk for AEs in nursing infants secondary to treatment of the parent with Mozobil™, breastfeeding should be discontinued if the breastfeeding parent will be treated with Mozobil™.

**Injection site reactions: A** variety of injection site reactions have been reported after injecting Mozobil™. While very common, most of these resolve within minutes with no treatment. Prolonged reactions if problematic will be treated with standard medical therapies such as topical corticosteroids, oral antihistamines, etc.

Musculoskeletal Pain: Musculoskeletal pain may affect bones, muscles, ligaments, tendons, or nerves and may include one or a combination of the following types of pain: bone pain (deep throbbing or pulsatile dull pain from the back, legs, and arms), joint pain or arthralgia/arthritis (pain and/or swelling in the joints), back pain, and or pain in the extremities.

**Gastrointestinal side effects:** Various side effects related to gastrointestinal function have been reported after doses of 0.24 mg/kg including abdominal pain, diarrhea, constipation, nausea, and vomiting, all at higher rates than in patients given placebo. In general, these have not resulted in the need to stop the medication.

**Excessively high white counts (leukocytosis):** Because of the mechanism of action of this medication, it is expected that WBC will rise. However, normal volunteers have not risen above 25,000 cells per microliter of blood after 1 day of treatment at 0.24 mg/kg/day. The patients likely to be enrolled in this study generally have low circulating WBC but also have more cells than normal sequestered in the bone marrow. This is why we have designed this study to start at lower drug levels that are slowly increased (dose escalation).

**Thrombocytopenia and bleeding:** Lower platelet counts (thrombocytopenia) have been reported with administration of this drug, but no serious bleeding events have been reported to date. We will closely monitor the platelet count 4X/day during the dose escalation phase, and the drug will be stopped if dangerously low platelet counts (<20,000/microliter of blood) are encountered.

**Splenic enlargement:** An increase in spleen size has been shown to occur in rats treated with the study agent at proportionately higher doses than will be used here. Although this has not been reported as causing any severe problems in humans, we will monitor for clinical signs of problems by asking patients about left shoulder or left upper quadrant abdominal pain each day (case report form). If these occur, the patient will be evaluated by medically appropriate imaging and the study drug may be stopped at the discretion of the PI.

**Cardiac Arrhythmia: In one human study, 2 HIV-infected patients developed a non‑life‑threatening cardiac arrhythmia consisting of frequent premature ventricular contractions. Individuals felt to be at higher risk for this problem based on EKG or cardiac problems detected on ECHO or who have a past history of cardiac arrhythmia will be excluded from this study.**

**Paresthesia*:* In one human study, some HIV-infected patients developed some transient perioral or peripheral numbness that resolved with discontinuation of the drug. The patients affected were treated with doses considerably higher than will be used in this study and the problem may have been related to other medications, which HIV-infected patients commonly use, or HIV itself. Participants in this study will be asked whether they are suffering from this symptom and the drug will be discontinued if Grade III/IV toxicity develops.**

**Infection:** It is not currently known whether Mozobil^TM^ could change the risk of infection. For the purposes of this study, any individual with signs or symptoms of active infection will not be allowed to participate, and individuals who develop an infection during this study will have the study drug discontinued.

#### Risks and Discomforts of Phlebotomy

Subjects will undergo repeat blood sampling on several occasions during the initial phase of the study and during the treatment phase. Because of the frequency of blood draws, we will place a Hep-Lock or other venous access device in each patient unless patients already have such devices. For research blood purposes, each venipuncture will be for 3 mL to 120 mL of blood in adults (not to exceed limits set forth by the Clinical Center Medical Administrative Series Policy #: M95-9: Guidelines for Limits of Blood Drawn for Research Purposes in the Clinical). Risks of phlebotomy and Hep-Lock placement include pain, ecchymosis, infection, bleeding at the insertion site, hematoma, and fainting.

#### Risks and Discomforts of Wart Biopsy

Wart biopsies may be requested for diagnosis and research from patients but will be optional. An experienced dermatologist will perform the procedure. Local anesthetics will only be used to prepare the biopsy site; no conscious sedation or general anesthesia will be given. The risks of wart biopsy include local pain, bleeding, infection, and the potential for scar and keloid formation. Antibiotics and oral analgesics will be used to manage pain and infection. The histologic response to Mozobil^TM^ offers generalizable knowledge about the pathophysiology and treatment of this condition.

#### Risks and Discomforts of Bone Marrow Biopsy

Patients will have the option of having bone marrow biopsy/aspiration done for research purposes, which could be done either as outpatient or inpatient. Local anesthetics will be used to prepare the biopsy site and IV conscious sedation may be offered to patients for comfort; these agents may result in transient local discomfort initially and occasionally be associated with allergic reactions. General anesthesia will only be given if the participant is admitted for the procedure. The primary risks of bone marrow biopsy include local pain, bleeding, and infection. Antibiotics and oral analgesics will be used to manage pain and infection.

#### Risks and Discomforts of HIV Testing

In addition to the usual risks and discomforts of phlebotomy, specific consent for HIV antibody testing will be obtained. The attendant responsibilities of reporting, notification of partners, and limits of confidentiality will be fully discussed with study subjects.

#### Risks of Stopping G-CSF/GM-CSF

In order to measure the effect of drug on ANC, we need to temporarily hold medications that could have an effect on white count, especially G-CSF or GM-CSF. These medications will be withheld for at least two days in advance of starting study medication and throughout this study. This could slightly increase the risk of infection; however, this risk is relatively small since the half-life of these agents is about one day meaning that the ANC levels will slowly drop to baseline once their administration is stopped. In normal volunteer studies, it has taken 5 days for the ANC to return to baseline after a single dose of G-CSF. These drugs will be restarted at the pre-study dose as soon as Active phase B of the study is completed unless patient elects to participate on Phase C or D. If there are any clinical indications of infection either before, during, or after the administration of the study drug in phase A or B, the study drug's administration will be halted. However, in Phase C and D, the response to infection will depend on the ANC achieved. If the patient’s nadir ANC is ≤250/µL at the time of the infection, the study drug will be stopped, and appropriate antibiotics and G-CSF will be administered; however, if the ANC is >250/µL, then the patient will continue Mozobil. Patients will be admitted to the NIH Clinical Center for 2 days prior to initial dosage of the study drug for observation, and appropriate screening for any infection will be performed during this period. As an additional safety feature, patients who are admitted on prophylactic antibiotics will be continued on these throughout the study.

#### Risk of Genetic Testing

In some instances, DNA will be extracted from tissue specimens and subjected to HLA testing or screened for gene mutations and gene dysfunction. The results of genetic testing may determine biological relation/status of patients to their legal parents, a fact that may not have been known previously. This may have possible effects on patient’s emotional well-being. Also if this information were released to patient, their family, or third parties, it could potentially be misused. Such misuse could lead to adverse psychological effects or undesired effects on the ability of the patient or their family members to obtain a job or insurance. In order to minimize these potential risks, all research information obtained from patient’s blood or tissue samples will be kept confidential and such DNA will be coded and maintained in a way that protects patients’ privacy. In some cases, coded DNA samples will be sent to other investigators for additional study. The PI on this protocol will prepare the final written report with clinical and genetic interpretations for both positive and negative sequencing results that will be sent to the referring physician upon patient’s consent.

#### Risks and Discomforts of the Infusion Pump Device

Patients require training to utilize the pump. The pump must remain attached continuously for three days and can be inadvertently dislodged. The adhesive may irritate the skin and the area around the cannula might rarely become infected. Patients must be proficient in setting-up, activating, dosing, monitoring, and deactivating the pump. They need ensure that they have their controller and a supply of pumps and plerixafor for replacement every three days, or more frequently if the pump is inadvertently dislodged.

### STUDY POTENTIAL BENEFIT

**Correction of Neutropenia:** The primary outcome of this study is the correction of neutropenia. Because of the important role of CXCR4 in myelokathexis in WHIMS and the rise in WBC and neutrophil numbers in normal volunteers, it is likely that the drug will increase neutrophil counts in participants. It is currently unknown what dose is effective and whether the rise will be to a clinically significant level and that is what this study is designed to begin to answer.

**Reduction of Risk of Infection:** Correction of neutropenia by release of mature and functional neutrophils from the bone marrow to the circulation is likely to reduce the risk of infection by allowing these white cells to move to sites of infection. Reduction in infection risk is not expected to be clinically significant unless participation in phase C or D is completed. An exploratory endpoint will be to compare an annualized infection risk on Mozobil to that on G‑CSF or before hematopoietic stimulation therapy.

**Correction of Hypogammaglobulinemia:** It is believed that the abnormal function of CXCR4 on mature B cells or plasma cells may cause the hypogammaglobulinemia seen in WHIMS. Therefore, it is possible that a reduction in the function of CXCR4 may help to correct this and allow a more durable response to vaccination. It is possible that during phases C and D, and especially with continuous infusion pump in Phase D that maintaining steady levels of circulating B and T cells may encourage normal development of mature B cells or plasma cells. Thus, we may see improvement of immunoglobulin levels or vaccine responses.

**Clearance of HPV Infection:** The susceptibility of individuals with WHIMS to HPV is poorly understood, but such infection could respond to drug treatment. However, this outcome is likely to require long-term administration of drug, which is not part of phase A/B of this study. It is possible that during phases C and D we may see improvement of HPV lesions, which is why we are photographing and biopsying these.

### HUMAN SUBJECT PROTECTIONS

#### Institutional Review Board

A copy of the protocol proposed informed consent form, other written subject information, and any proposed advertising material will be submitted to the IRB for written approval.

The investigator must submit and, where necessary, obtain approval from the IRB for all subsequent protocol amendments and changes to the informed consent document. The investigator will notify the IRB of deviations from the protocol and SAEs.

The investigator will be responsible for obtaining IRB approval of the annual Continuing Review throughout the duration of the study.

#### Protocol Amendments and Study Termination

No revisions to this protocol will be permitted without documented approval from the IRB that granted the original approval for the study. This does not apply to changes made to reduce discomfort or avert risk to study volunteers. Furthermore, in the event of a medical emergency, the investigators shall perform any medical procedures that are deemed medically appropriate. The PI must notify the IRB of all such occurrences.

The PI and NIAID reserve the right to terminate the study. The PI will notify the IRB in writing of the study’s completion or early termination.

#### Privacy and Confidentiality Provisions

The privacy and confidentiality of the participating subjects will be protected to the extent required by federal, state, and local law. The investigator will ensure that the subject’s anonymity is maintained. Subjects will not be identified in any publicly released reports of this study. All records will be kept confidential to the extent required by federal, state, and local law. The study monitors may inspect all documents and records required to be maintained by the Investigator, including but not limited to, medical records. The investigator will inform the subjects that the above-named representatives will review their study-related records without violating the confidentiality of the subjects. All laboratory specimens, evaluation forms, reports, and other records that leave the site will be identified only by a coded number in order to maintain subject confidentiality. All records will be kept locked and all computer entry and networking programs will be done with coded numbers only. Clinical information will not be released without written permission of the subject, except as necessary for monitoring by IRB, NIAID, or OHRP.

To further protect the privacy of study participants, a Certificate of Confidentiality has been issued by the NIH. This certificate protects identifiable research information from forced disclosure. It allows the investigator and others who have access to research records to refuse to disclose identifying information on research participation in any civil, criminal, administrative, legislative, or other proceeding, whether at the federal, state, or local level. By protecting researchers and institutions from being compelled to disclose information that would identify research participants, Certificates of Confidentiality help achieve the research objectives and promote participation in studies by helping assure confidentiality and privacy to participants.

#### Considerations of Alternatives to Participation

Current therapeutic recommendations based on limited clinical experience and no controlled clinical trials in WHIMS patients involve daily or every other day injection of G-CSF to normalize neutrophil counts and monthly infusion of IVIG.[7] These therapies are expensive, non-specific, have significant side effects and toxicities, and do not fully correct the patient’s problems, especially warts and HPV-related cancers. These options will be presented to patients, but will be administered by the primary care provider.

#### Policy Regarding Research-Related Injuries

The Clinical Center will provide short-term medical care for any injury resulting from participation in this research. In general, the NIH, the Clinical Center, or the Federal Government will provide no long-term medical care or financial compensation for research-related injuries.

#### Subject Remuneration

Study subjects and their families will not receive any per visit reimbursement. However, we will provide transportation costs to and from the NIH for all study participants if financial assistance is required as assessed by an NIH social worker. Patient who agrees to the optional wart biopsies will be compensated a total of $120.00 per biopsy. Patients who agree to optional bone marrow biopsies will be compensated a total of $200.00 per biopsy.

### SAFETY ASSESSMENT

AEs and other reportable events are defined in NIH Policy 801: Reporting Research Events.

#### Reporting Procedures

Unanticipated problems, non-compliance, and other reportable events will be reported to the NIH IRB according to Policy 801.

#### Reporting of Pregnancy

Pregnancy itself is not an AE. However, complications from pregnancy are AEs and may be SAEs. Pertinent obstetrical information for all pregnancies will be reported to the NIAID Clinical Safety Office (CSO) via fax or e-mail within 5 business days from the site’s awareness of the pregnancy.

Pregnancy outcome data (e.g., delivery outcome, spontaneous, or elective termination of the pregnancy) will be reported to the CSO within 5 business days of the site’s awareness of the outcome on a protocol-specified form. In the event of pregnancy, the subject will be followed to term for safety monitoring. In the event of pregnancy:

- If patient is being treated with plerixafor, the study drug will be discontinued;
- If patient is being treated with G-CSF, the study comparator drug may be withheld for 1^st^ trimester;
- Patient will be advised to notify their obstetrician;

Pregnancy will be reported to the Safety Monitoring Committee (SMC).

#### Toxicity Criteria

The severity of AEs will be assessed according to the relevant sections of the National Cancer Institute (NCI) Common Toxicity Criteria Scale (CTC version 3.0 [<http://ctep.cancer.gov/protocolDevelopment/electronic_applications/ctc.htm> (please see APPENDIX A). The following definitions will be used for toxicities that are not defined in the Common Toxicity Criteria Scale (please note that dose adjustment of the study drug is not applicable to this study):

1. Mild (Grade 1): The AE is noticeable to the patient but does not interfere with routine activity. The AE does not require discontinuing the study procedures.
2. Moderate (Grade 2): The AE interferes with routine activity but responds to symptomatic therapy or rest. The AE may require modifying the study procedures but not discontinuing the study procedures.
3. Severe (Grade 3): The AE significantly limits the subject’s ability to perform routine activities despite symptomatic therapy. In addition, the AE leads to discontinuing the study procedures.
4. Life-threatening (Grade 4): The AE requires discontinuing the study procedures. The patient is at immediate risk of death.
5. Death related to SAE (Grade 5).

Abnormal biological or vital sign values that are considered clinically relevant by the investigator will be reported as an AE or as an SAE. Certain information, while not necessarily meeting the definition of an AE, may nonetheless be of value for reporting.

### DATA AND SAFETY MONITORING

The data generated during this study will be monitored by the PI for safety and compliance with protocol-specified requirements. The trial will be conducted in compliance with this protocol, International Council on Harmonisation of Technical Requirements for Pharmaceuticals for Human Use (ICH) GCP, and any applicable regulatory requirement(s). Close cooperation between the designated members of the study team will occur to evaluate and respond to individual AE in a timely manner.

A Grade 3 or 4 SAE occurrence that is possibly, probably, or definitely related to study drug in any particular participant will necessitate the protocol being placed on voluntary hold by the PI until the safety data have undergone a review by the IRB. In the event of an emergency outside the facilities of NIH, the home physician will assess the situation and liaise with the PI accordingly, and transfer of care to NIH can be organized as clinically indicated.

All privacy and confidentiality of all participants will be maintained in accordance with NIH guidelines and policy.

### PROTOCOL MONITORING PLAN

Since this protocol carries more than a minimum risk, we will establish an SMC within NIAID/LMI. The goal of this SMC will be to ensure patient safety and study integrity. This committee will be comprised of three independent experts in the field of inherited immunodeficiency and severe congenital neutropenia. The independent experts do not have direct involvement in the conduct of the study. The SMC will meet at least every 6 months (depending on enrollment) with the minimal presence of two SMC specialists, PI, and (2) AIs. Please see APPENDIX E for SMC plan.

### PLAN FOR USE AND STORAGE OF BIOLOGICAL SAMPLES

#### Use of Stored Samples and Data

Sample collection, handling, storage, and shipment will follow Clinical Center policies and procedures. We will make a summary of radiology and laboratory results available to patients and their private physicians.

Samples and data collected under this protocol may be used to study the causes of primary immunodeficiency, the consequences thereof, and its treatment. Appropriate genetic testing to determine the cause of the underlying disorder will be performed unless this has already been done prior to study entry. This testing may have the unintended consequence of identifying non-paternity or increased risk of other disorders.

#### Disposition of Stored Samples and Data

The following criteria apply to all stored samples and data:

- Access to stored samples will be limited using either a locked room or a locked freezer. Samples and data will be stored using codes assigned by the investigators. Data will be kept in password-protected computers. Only investigators will have access to the samples and data.
- Samples and data acquired will be tracked using software designed for this purpose.
- Any loss or unanticipated destruction of samples or data (for example, due to freezer malfunction) that meets the definition of protocol deviation and/or compromises the scientific integrity of the data collected for the study; will be reported to the NIH IRB.
- At the completion of the protocol (termination), samples may be kept for future research.
- Additionally, subjects may decide at any point not to have their samples stored. In this case, the PI will destroy or assure the destruction of all known remaining samples and report what was done to both the subject and to the IRB. This decision will not affect the subject’s participation in other protocols at NIH.

### PUBLICATION POLICY

Following completion of the study, the investigator may publish the results of this research in a scientific journal. The International Committee of Medical Journal Editors (ICMJE) member journals have adopted a trials-registration policy as a condition for publication. This policy requires that all clinical trials be registered in a public trials registry such as [ClinicalTrials.gov](http://prsinfo.clinicaltrials.gov/), which is sponsored by the National Library of Medicine. Other biomedical journals are considering adopting similar policies. It is the responsibility of the NIAID Division or Branch to register this trial in an acceptable registry. Any clinical trial starting enrollment after 01 July 2005 must be registered either on or before the onset of patient enrollment.

### REFERENCES

1.	Ward, A.C. and D.C. Dale, *Genetic and molecular diagnosis of severe congenital neutropenia.* Curr Opin Hematol, 2009. **16**(1): p. 9-13.

2.	Dale, D.C., et al., *The Severe Chronic Neutropenia International Registry: 10-Year Follow-up Report.* Support Cancer Ther, 2006. **3**(4): p. 220-31.

3.	Kawai, T., et al., *WHIM syndrome myelokathexis reproduced in the NOD/SCID mouse xenotransplant model engrafted with healthy human stem cells transduced with C-terminus-truncated CXCR4.* Blood, 2007. **109**(1): p. 78-84.

4.	Kawai, T., et al., *Enhanced function with decreased internalization of carboxy-terminus truncated CXCR4 responsible for WHIM syndrome.* Exp Hematol, 2005. **33**(4): p. 460-8.

5.	Hernandez, P.A., et al., *Mutations in the chemokine receptor gene CXCR4 are associated with WHIM syndrome, a combined immunodeficiency disease.* Nat Genet, 2003. **34**(1): p. 70-4.

6.	Diaz, G.A., *CXCR4 mutations in WHIM syndrome: a misguided immune system?* Immunol Rev, 2005. **203**: p. 235-43.

7.	Kawai, T. and H.L. Malech, *WHIM syndrome: congenital immune deficiency disease.* Curr Opin Hematol, 2009. **16**(1): p. 20-6.

8.	Balabanian, K., et al., *Leukocyte analysis from WHIM syndrome patients reveals a pivotal role for GRK3 in CXCR4 signaling.* J Clin Invest, 2008. **118**(3): p. 1074-84.

9.	Chae, K.M., J.O. Ertle, and M.D. Tharp, *B-cell lymphoma in a patient with WHIM syndrome.* J Am Acad Dermatol, 2001. **44**(1): p. 124-8.

10.	Imashuku, S., et al., *Epstein-Barr virus-associated T-lymphoproliferative disease with hemophagocytic syndrome, followed by fatal intestinal B lymphoma in a young adult female with WHIM syndrome. Warts, hypogammaglobulinemia, infections, and myelokathexis.* Ann Hematol, 2002. **81**(8): p. 470-3.

11.	Dale, D.C., et al., *The CXCR4 antagonist plerixafor is a potential therapy for myelokathexis, WHIM syndrome.* Blood, 2011.

12.	McDermott, D.H., et al., *The CXCR4 antagonist plerixafor corrects panleukopenia in patients with WHIM syndrome.* Blood, 2011. **118**(18): p. 4957-62.

13.	McDermott, D.H., et al., *A phase 1 clinical trial of long-term, low-dose treatment of WHIM syndrome with the CXCR4 antagonist plerixafor.* Blood, 2014. **123**(15): p. 2308-2316.

14.	McDermott, D.H., et al., *Plerixafor for the Treatment of WHIM Syndrome.* N Engl J Med, 2019. **380**(2): p. 163-170.

15.	McDermott, D.H., et al., *A phase III randomized crossover trial of plerixafor versus G-CSF for treatment of WHIM syndrome.* J Clin Invest, 2023. **133**(19).

16.	Winer, K.K., et al., *Effects of pump versus twice-daily injection delivery of synthetic parathyroid hormone 1-34 in children with severe congenital hypoparathyroidism.* J Pediatr, 2014. **165**(3): p. 556-63 e1.

17.	Polonsky, W.H., et al., *Impact of the Omnipod((R)) Insulin Management System on Quality of Life: A Survey of Current Users.* Diabetes Technol Ther, 2016. **18**(10): p. 664-670.

18.	De Clercq, E., *The bicyclam AMD3100 story.* Nat Rev Drug Discov, 2003. **2**(7): p. 581-7.

19.	Donzella, G.A., et al., *AMD3100, a small molecule inhibitor of HIV-1 entry via the CXCR4 co-receptor.* Nat Med, 1998. **4**(1): p. 72-7.

20.	Lack, N.A., et al., *A pharmacokinetic-pharmacodynamic model for the mobilization of CD34+ hematopoietic progenitor cells by AMD3100.* Clin Pharmacol Ther, 2005. **77**(5): p. 427-36.

21.	Hendrix, C.W., et al., *Safety, pharmacokinetics, and antiviral activity of AMD3100, a selective CXCR4 receptor inhibitor, in HIV-1 infection.* J Acquir Immune Defic Syndr, 2004. **37**(2): p. 1253-62.

22.	Nagasawa, T., et al., *Defects of B-cell lymphopoiesis and bone-marrow myelopoiesis in mice lacking the CXC chemokine PBSF/SDF-1.* Nature, 1996. **382**(6592): p. 635-8.

23.	Zou, Y.R., et al., *Function of the chemokine receptor CXCR4 in haematopoiesis and in cerebellar development.* Nature, 1998. **393**(6685): p. 595-9.

24.	Ma, Q., et al., *Impaired B-lymphopoiesis, myelopoiesis, and derailed cerebellar neuron migration in CXCR4- and SDF-1-deficient mice.* Proc Natl Acad Sci U S A, 1998. **95**(16): p. 9448-53.

25.	Rao, G.V.N., et al., *Ultra performance liquid chromatographic method for simultaneous quantification of plerixafor and related substances in an injection formulation.* Cogent Chemistry, 2017. **3**.

26.	Seki, J.T., et al., *Chemical Stability of Plerixafor after Opening of Single-Use Vial.* Canadian Journal of Hospital Pharmacy, 2017. **70**(4): p. 270-275.

27.	Kim, S.H., J. Thiesen, and I. Kramer, *Physiochemical Stability of Mozobil(R) (Plerixafor) Solution for Injection in Glass Vials and Plastic Syringes over a Three-Month Storage Period.* Pharm. Technol. Hosp. Pharm., 2016. **1**(2): p. 73-81.

APPENDIX A: TOXICITY TABLE

| **ALLERGY/IMMUNOLOGY** | | | | | | | |
| --- | --- | --- | --- | --- | --- | --- | --- |
|  | | | GRADE | | | | |
| **Adverse Event** | **Short Name** | | **1** | **2** | **3** | **4** | **5** |
| Allergic reaction/  hypersensitivity  (including drug fever) | Allergic reaction | | Transient flushing or  rash; drug fever <38°C  (<100.4°F) | Rash; flushing; urticaria;  dyspnea; drug fever  ≥38°C ("100.4°F) | Symptomatic  bronchospasm, with or  without urticaria;  parenteral medication(s)  indicated; allergy-related edema/ angioedema;  hypotension | Anaphylaxis | Death |
| Autoimmune reaction | Autoimmune reaction | | Asymptomatic and serologic or other evidence of autoimmune  reaction, with normal organ function and  intervention not indicated | Evidence of autoimmune  reaction involving a nonessential organ or function (e.g.,  hypothyroidism) | Reversible autoimmune  reaction involving function  of a major organ or other adverse event (e.g., transient colitis or anemia) | Autoimmune reaction with  life-threatening  consequences | Death |
| **CONSTITUTIONAL SYMPTOMS** | | | | | | | |
| Fatigue  (asthenia, lethargy,  malaise) | | Fatigue | Mild fatigue over baseline | Moderate or causing  difficulty performing some ADL | Severe fatigue interfering  with ADL | Disabling | --- |
| Insomnia | | Insomnia | Occasional difficulty  sleeping, not interfering  with function | Difficulty sleeping,  interfering with function but not interfering with ADL | Frequent difficulty sleeping, interfering with  ADL | Disabling | ---- |
| **DEATH** | | | | | | | |
| Death not associated with Multi-organ failure  – Sudden death | | Death not associated with Multi-organ failure  – Sudden death | ---- | ---- | ----- | ----- | ---- |
| **DERMATOLOGY/SKIN** | | | | | | | |
| Injection site reaction/  extravasation changes | | Injection site reaction | Pain; itching; erythema | Pain or swelling, with inflammation or phlebitis | Ulceration or necrosis that is severe; operative  intervention indicated | ------- | ------ |
| Urticaria  (hives, welts, wheals) | | Urticaria | Intervention not indicated | Intervention indicated for <24 hrs | Intervention indicated for  ≥24 hrs | ------ |  |
| **GASTROINTESTINAL** | | | | | | | |
| Diarrhea | | Diarrhea | Increase of <4 stools per  day over baseline; mild  increase in ostomy output  compared to baseline | Increase of 4 – 6 stools  per day over baseline; IV fluids indicated <24hrs;  moderate increase in  ostomy output compared to baseline; not interfering with ADL | Increase of "7 stools per  day over baseline;  incontinence; IV fluids>24 hrs; hospitalization;  severe increase in  ostomy output compared to baseline; interfering  with ADL | Life-threatening  consequences (e.g.,  hemodynamic collapse) | Death |
| Flatulence | | Flatulence | Mild | Moderate | ------ | ----- | ------- |
| Nausea | | Nausea | Loss of appetite without alteration in eating habits | Oral intake decreased  without significant weight loss, dehydration or  malnutrition; IV fluids  indicated <24 hrs | Inadequate oral caloric or  fluid intake; IV fluids, tube  feedings, or TPN  indicated >24 hrs | Life-threatening  consequences | Death |
| Vomiting | | Vomiting | 1 episode in 24 hrs | 2 – 5 episodes in 24 hrs;  IV fluids indicated  <24 hrs | ≥6 episodes in 24 hrs; IV  fluids, or TPN indicated  ≥24 hrs | Life-threatening  consequences | Death |
| **MUSCULOSKELETAL/SOFT TISSUE** | | | | | | | |
| Arthralgia | | Arthralgia | Mild | Moderate | Severe | Disabling | --- |
| **NEUROLOGY** | | | | | | | |
| Dizziness | | Dizziness | With head movements or  nystagmus only; not  interfering with function | Interfering with function,  but not interfering with  ADL | Interfering with ADL | Disabling |  |
| Headache | | Headache | Mild headache not interfering  with function | Moderate headache; pain or  analgesics interfering with  function, but not  interfering with ADL | Severe headache; pain or  analgesics severely  interfering with ADL | Disabling |  |
| **INFECTION** | | | | | | | |
| Infection  (documented clinically or  microbiologically) | | Infection | ----------- | Localized, local  intervention indicated | IV antibiotic, antifungal, or  antiviral intervention  indicated; interventional  radiology or operative  intervention indicated | Life-threatening  consequences (e.g.,  septic shock,  hypotension, acidosis,  necrosis) | Death |
| **PAIN** | | | | | | | |
| Pain associated with Splenic enlargement | | Pain associated with Splenic enlargement | Mild pain not interfering with function | Moderate pain; pain or analgesics interfering with function but not interfering with ADL | Severe pain; pain or analgesics severely interfering with ADL | Disabling | ----- |
| **BLOOD/BONE MARROW** | | | | | | | |
| Platelets | | Platelets | <LLN – 75,000/mm3  <LLN – 75.0 x 109 /L | <75,000 – 50,000/mm3  <75.0 – 50.0 x 109 /L | <50,000 – 20,000/mm3  <50.0 – 25.0 x 109 /L | <20,000/mm3  <20.0 x 109 /L | Death |

APPENDIX B: SCHEDULE OF PROCEDURES/EVALUATIONS for Phase A and/or B

**Prior to Initiating Drug:**

- Ensure patient meets eligibility requirements and has no exclusion criteria.
- Obtain informed consent for protocol.
- Make sure that patient has been off G-CSF or GM-CSF for at least 2 days (washout period).
- Make sure safety laboratory tests, an electrocardiogram, and an echocardiogram have been done and do not reveal congenital or other heart disease that would significantly raise the risk of arrhythmia.
- Make sure that there is a CBC documented in the chart that shows neutropenia (ANC<1000 cells/microliter of blood).
- Ensure that the patient is not currently infected by a standard review of systems and physical examination and cultures if clinically indicated.
- Record patient's height, weight, and vital signs.
- Determine baseline CBC, chemistries, immunoglobulin levels, complement levels, B cell immunophenotyping panels I and II, and any research laboratory tests.
- Admit patient to the NIH Clinical Research Center for frequent blood draws and vital signs.
- Administer an initial dose of 0.02 mg/kg of actual body weight of Mozobil™ sq qd if CrCl is >50 mL/min calculated by Cockroft-Gault formula and obtain CBC profiles prior to each dose. If CrCl is <50mL/min, then decrease starting dose by 1/3.

**After Initiating Drug:**

- Following schedule above in Section 4.2.2, increase dose each day until absolute neutrophil count is >4000 cells/microliter. Total daily dose not to exceed 40 mg for CrCl >50 mL/min or 27 mg for CrCl <50 mL/min and not to exceed 0.24 mg/kg/dose for CrCl >50 mL/min and 0.16 mg/kg/dose for CrCl <50 mL/min. Once the absolute neutrophil count reaches 4000 cells/microliter, no further dose escalation will be given. No sooner than 48 hours later, we will start Phase B (pharmacokinetic monitoring). At this phase, we will give the same dose that achieved the absolute neutrophil count of 4000 cells/microliter or the FDA approved dose if phase A not performed first and blood will be obtained at set intervals for pharmacokinetic and leukocyte response purposes.
- Closely monitor the patient for rare systemic side effects such as vasovagal or anaphylactoid reactions for 1 hour after each dose. Monitor renal function and platelet count at least daily. Treat gastrointestinal or skin site injection reactions as medically indicated.

APPENDIX C: DOSING OF MOZOBIL^TM^ IF THERE IS RENAL IMPAIRMENT

In patients with moderate and severe renal impairment (estimated creatinine clearance [CrCl] ≤50 mL/min), reduce each dose of Mozobil^TM^ in the dose escalation by one-third (maximum dose 0.16 mg/kg/day) as shown in Table 1 (below). If CrCl is ≤50 mL/min, the dose should not exceed 27 mg/day. Because of the renal excretion of the drug, this reduction in patients with moderate and severe renal impairment is expected to result in similar drug levels compared with subjects with normal renal function.

| Table 1: Recommended Dosage of Plerixafor in Patients with Renal Impairment | |
| --- | --- |
| Estimated Creatinine Clearance (mL/min) | Dose |
| >50 | 0.24 mg/kg/day (not to exceed 40 mg/day) |
| ≤50 | 0.16 mg/kg/day (not to exceed 27 mg/day) |

The following (Cockroft-Gault) formula may be used to estimate Creatinine Clearance (CrCl):

    Males:
    Creatinine clearance (mL/min) = weight (kg) X (140 – age in years)
                                      72 X serum creatinine (mg/dL)

    Females:
    Creatinine clearance (mL/min) = 0.85 X value calculated for males

Since there is insufficient information to make dosage recommendations in patients on hemodialysis, these individuals are excluded from this study.

APPENDIX D: FDA APPROVED PACKAGE INSERT FOR MOZOBIL^TM^

Please see the attached document.

APPENDIX E: NIAID/LMI SAFETY MONITORING COMMITTEE

Protocol Title: A Phase I Study of Mozobil^TM^ in the Treatment of Patients with WHIMS

**Mission of the NIAID/LMI Safety Monitoring Committee**

The purpose of this study is to assess the safety and efficacy of Mozobil™ in the treatment of neutropenia resulting from the abnormal retention of mature neutrophils and other white blood cells in the bone marrow (myelokathexis) in WHIMS patients.

Since this protocol carries more than a minimum risk, we will establish a Safety Monitoring Committee (SMC) within NIAID/LMI. The goal of this SMC will be to ensure patient safety and study integrity. At each meeting, the SMC will review the following:

- All notable events (including SAE’s and AE line listings).
- NIH interim clinical laboratory data
- All enrollment and withdrawal from participation data
- Summary of study data.

**MEMBERS OF THE SAFETY MONITORING COMMITTEE**

This committee will be comprised of three independent experts in the field of inherited immunodeficiency and severe congenital neutropenia. The independent experts do not have direct involvement in the conduct of this study and have no other interests with any of the collaborating or competing pharmaceutical firms who manufacture similar drugs. To ensure this, we will obtain DEC clearance of all independent experts. Although the experts are employed by NIH/NIAID, none are in a supervisory role of the named investigators (PI and AI) involved in this trial.

Specialists (independent expert reviewers)

Michail Lionakis, MD (Infectious Disease)	NIAID/LCIM	301-443-5089

Brian Kelsall, MD (Immunology)	``	NIAID/LMI	301-496-8493

Josh Farber, MD (Immunology)	NIAID/LMI	301-402-4910

**RESEARCH TEAM PARTICIPANTS**

Research team members may participate in SMC reviews. The research team participants will not have voting privileges.

**FREQUENCY OF MEETINGS**

The SMC will meet at least every 6 months (if there are new enrollments not yet reviewed that have completed the drug administration phase) with the minimal presence of 2 SMC specialists, PI, (2) Associate Investigators. In the event that these criteria for a valid meeting cannot be met, then the meeting will be convened on a different day within 2 weeks before or after the regularly scheduled time. In addition, the SMC has the authority to request additional SMC review meetings as they deem necessary.

**VOTING**

At the end of each meeting, voting members (3 independent experts) will discuss and vote on the recommendations. A quorum, defined as a majority of 3 independent experts, is required for voting.

**MEETING NOTES AND COMMITTEE ACTIONS**

Study Coordinator will provide administrative support to schedule review meetings, distribute review materials, draft meeting summary reports and distribute final reports to appropriate individuals. The attached NIH Policy for Data and Safety Monitoring policy will be followed for the duration of the study. Final SMC reports will be kept in the regulatory binder and submitted as part of continuing review to NIH IRB. If, however, there is as an SMC recommendation to hold enrollment, further treatment, and/or modify the protocol or consent(s), the PI will send the SMC recommendations to the NIH IRB directly, together with a formal amendment to the protocol or consent.

APPENDIX F: SCHEDULE OF EVENTS FOR PHASES C AND D

| **Evaluations** | **Screening and Baseline** | | **Start of Drug and Dose Adjustment** | | | **Treatment Period**  **(Phase D max is 24 months)** | | | **End of Treatment** | |
| --- | --- | --- | --- | --- | --- | --- | --- | --- | --- | --- |
|  | *Screen* | *Baseline* | *Washout*  D -2 | *Start Infusion*  D 0-2 | Adjust Dose  M 1-3 | Q3 months | Q6 months | *Annual* | End of Treatment  (Phase D @ 12M) | End of Study  6 ±2M |
| Informed consent (Phase D), ECHO (within 5 years), PT/PTT, Viral Panel *^a^*. Training: Memory Aid & Infusion Pump (w/ saline). | X | X |  |  |  |  |  |  |  |  |
| Ultrasound spleen, EKG, Urinalysis |  | X |  |  |  |  |  | X |  |  |
| Home Infusion Checklist, 2-day Washout. |  |  | X |  |  |  |  |  |  |  |
| H&P, Vital Signs, Memory Aid, Infection & AE review, Infusion data, Contraceptive counseling, Photos, Pregnancy test, CRP, ESR, Immunoglobulins. Research: B12, Folate, Blood, Antibody titers, HPV swabs. |  | X |  |  |  |  | X | X | X | X |
| WHIM Annual Consults: Dermatology, Dental, GYN, ENT, Audiology, PFT. |  | X |  |  |  |  |  | X |  |  |
| CBC, TBNK, CRP, Chemistry *^b^* |  | X |  | X ^c^ | L ^d^ | L ^d^ | X | X | X | X |
| Wart, skin, & bone marrow biopsies. |  | O |  |  |  |  |  | O | O |  |

V=visit; D=study Day; M= Study Month; O=Optional; L= may be performed at subject’s Local facility.

CBC=complete blood count; ESR; PT/PTT=prothrombin time/partial thromboplastin time; EKG=electrocardiogram; ENT=ear, nose, and throat; AEs=adverse events.

^a^ Includes HIV, hepatitis B/C. HTLV type 1 and II.

^b^ Chemistry: NIH -Acute Care Panel, Hepatic Panel, Mineral Panel, Serum Protein, Lactate Dehydrogenase, Serum Uric Acid, & Creatine Kinase, and ESR. For home monitoring (local labs), BUN and creatinine are required. Additional chemistries labs may be requested, and CRP if available may be requested.

^c^ Chemistry and CRP not required. However, up to 4 CBCs and/or TBNK may be drawn each day to determine leukocyte response.

^d^ Local labs: BUN and Creatinine for Chemistry, CRP not required. However, CBCs and/or TBNK (if available) may be drawn to determine leukocyte response and for adjusting the dose. Monthly or as required to adjust dose for months 1-3.

APPENDIX G: A SAMPLE OF PLERIXAFOR DOSES, INFUSION RATES, AND STEADY STATE CONCENTRATIONS

| ***Weight*** | ***Dose*** | ***Infusion*** | | | ***Pump Daily*** | ***Pump Reservoir*** | ***Clearance ^[12]^*** | ***Concentration SS*** |
| --- | --- | --- | --- | --- | --- | --- | --- | --- |
| ***kg*** | ***mg/kg/day*** | ***mg/hr*** | ***mL/hr*** | ***Units/hr*** | ***mL / day*** | ***mL / 3.25 days*** | ***L / h*** | ***ng / mL*** |
| 70 | 0.01 | 0.0292 | 0.0015 | 0.15 | 0.0350 | 0.114 | 5.2 | 5.6 |
| 70 | 0.02 | 0.0583 | 0.0029 | 0.29 | 0.0700 | 0.228 |  | 11.2 |
| 70 | 0.03 | 0.0875 | 0.0044 | 0.44 | 0.1050 | 0.341 |  | 16.8 |
| 70 | 0.04 | 0.1167 | 0.0058 | 0.58 | 0.1400 | 0.455 |  | 22.5 |
| 70 | 0.05 | 0.1458 | 0.0073 | 0.73 | 0.1750 | 0.569 |  | 28.1 |
| 70 | 0.06 | 0.1750 | 0.0088 | 0.88 | 0.2100 | 0.683 |  | 33.7 |
| 70 | 0.07 | 0.2042 | 0.0102 | 1.02 | 0.2450 | 0.796 |  | 39.3 |
| 70 | 0.08 | 0.2333 | 0.0117 | 1.17 | 0.2800 | 0.910 |  | 44.9 |
| ***Weight*** | ***Dose*** | ***Infusion*** | | | ***Pump Daily*** | ***Pump Reservoir*** | ***Clearance ^[12]^*** | ***Concentration SS*** |
| ***kg*** | ***mg/kg/d*** | ***mg/hr*** | ***mL/hr*** | ***Units/hr*** | ***mL / day*** | ***mL / 3.25 days*** | ***L / h*** | ***ng / mL*** |
| 100 | 0.01 | 0.0417 | 0.0021 | 0.21 | 0.0500 | 0.163 | 7.4 | 5.6 |
| 100 | 0.02 | 0.0833 | 0.0042 | 0.42 | 0.1000 | 0.325 |  | 11.2 |
| 100 | 0.03 | 0.1250 | 0.0063 | 0.63 | 0.1500 | 0.488 |  | 16.8 |
| 100 | 0.04 | 0.1667 | 0.0083 | 0.83 | 0.2000 | 0.650 |  | 22.5 |
| 100 | 0.05 | 0.2083 | 0.0104 | 1.04 | 0.2500 | 0.813 |  | 28.1 |
| 100 | 0.06 | 0.2500 | 0.0125 | 1.25 | 0.3000 | 0.975 |  | 33.7 |
| 100 | 0.07 | 0.2917 | 0.0146 | 1.46 | 0.3500 | 1.138 |  | 39.3 |
| 100 | 0.08 | 0.3333 | 0.0167 | 1.67 | 0.4000 | 1.300 |  | 44.9 |

APPENDIX H: PLERIXAFOR INFUSION

| *Timeline:* | ***Day -2*** | ***Day -1*** | ***Day 0*** | ***Day 1*** | ***Day 2*** | ***Day 3*** | ***Weeks 2 & 4*** | ***Month 3*** | ***Month 12*** |
| --- | --- | --- | --- | --- | --- | --- | --- | --- | --- |
| *Activity:* |  | ***Training*** | ***Plerixafor Bolus*** | ***Plerixafor Infusion*** | ***NIH Dose Adj*** | ***NIH Dose Adj*** | ***Dose Adj*** |  |  |
| *Location:* | ***Optional NIH*** | ***NIH*** | ***NIH*** | ***NIH*** | ***NIH*** | ***Optional NIH*** | ***Home*** | ***Optional NIH*** | ***NIH*** |
| ***Pump (every 3 days)*** |  | Saline | Plerixafor #1 |  | Plerixafor #1 or 2 | Plerixafor #2 |  |  |  |
| ***Study Drug*** | Washout D2 | Washout D3 | Plerixafor Bolus | Plerixafor Continuous |  |  |  |  |  |
| ***Chemistry (chem 23)*** |  |  | 0h |  |  | Anytime | Anytime | Anytime | Anytime |
| ***CBC ^a^*** |  | 0, 3, 6h | 0/3/6/9h | 0/3/6/9h | > 24h | > 24h | >24h | >24h | >24h |
| ***TBNK (optional) ^a^*** |  |  | 3h | 3/6h | > 24h | > 24h | >24h | >24h | >24h |
| ***Lymphocyte Flow (NIH) ^a^*** |  |  | 0h |  |  |  |  | >24h | >24h |
| ***Antibody Titers (NIH)*** | Prior to Plerixafor | |  |  |  |  |  | Anytime | Anytime |
| ***Plerixafor Equivalent Dose*** |  |  | 0.015 mg/kg bolus | 0.03 mg/kg/d infusion | 0.01-0.08 mg/kg/d infusion | 0.01-0.08 mg/kg/d infusion | 0.01-0.08 mg/kg/d infusion | 0.01-0.08 mg/kg/d infusion | 0.01-0.08 mg/kg/d infusion |
| ***Review Infusion Hx*** |  | Yes | Yes | Yes | Yes | Yes |  | Yes | Yes |
| ***Target ANC*** |  |  |  | > 500 @ >24h | > 500 | > 500 | > 500 | > 500 | > 500 |
| ***Target ALC*** |  |  |  | > 1000 | > 1000 | > 1000 | > 1000 | > 1000 | > 1000 |
| ***Safety ANC ^c^*** |  |  |  |  |  | > 2x Baseline |  | Nadir > 250 by Month 3 | Nadir Average > 250 |

***^a^*** *“>” indicates lab should be drawn at steady state, for example “> 24h” means anytime greater than 24 hours after a change of dose or pump. 24 hours is the minimum wait time for measuring at steady state levels and assumes that the time between pump change is minimal (e.g. < 30 minutes) or the start of an infusion includes a loading dose. If practical or if in doubt, draw at > 24 hours after any change for a blood measurement to ensure drug serum level has reached steady state.*

*^c^****^c^*** *Subjects must achieve at least one measure of Absolute Neutrophil Count (ANC) of at least 250 per microliter and at least a two-fold increase in their Baseline ANC.*
